# Impact of in utero exposure to SARS-CoV-2 on children’s hospital admission: National birth cohort study of 262,000 children in England

**DOI:** 10.1101/2025.08.29.25333365

**Authors:** Mengyun Liu, Fariyo Abdullahi, Charlotte Jackson, Intira Jeannie Collins, Claire Thorne, Pia Hardelid

## Abstract

**Objective:** To examine the impact of in utero exposure to SARS-CoV-2 virus on children’s emergency and planned hospital admissions up to 40 months of age.

**Design, Setting, and Participants:** Nationwide, birth cohort study using multiple linked administrative datasets in England. Children with conception start dates between 1^st^ February and 31^st^ July 2020 and born alive to mothers residing in England, followed for 40 months.

**Exposure:** In utero SARS-CoV-2 exposure (i.e., maternal infection in pregnancy) during wild-type or Alpha variant dominant periods, characterised via linkage to national testing and hospital data.

**Main Outcome Measures:** First emergency or planned admission before 40 months of age.

**Results:** Of the 262,086 children included, 14,717 (5.6%) were exposed to SARS-CoV-2 in utero: 6,888 (2.6%) during the wild-type period and 7,829 (3.0%) during the Alpha period. Overall, 89,189 children (34.0%) had at least one emergency admission and 24,635 (9.4%) had at least one planned admission. After adjusting for socio-demographic characteristics and maternal chronic conditions, compared to children unexposed during either period, children exposed in utero during the wild-type period showed no difference in time to the first emergency admission, while Alpha-period exposure was associated with an 8% lower hazard (adjusted HR: 0.92, 99% CI: 0.87 to 0.97). No difference was found in the time to the first planned admission between in utero exposed groups and the unexposed group, nor in the time to either the first emergency or planned admission based on the trimester of in utero exposure.

**Conclusions:** In this large national birth cohort in England, we did not find an increased hazard of hospital admission by 40 months of age associated with in utero exposure to SARS-CoV-2, in the absence of maternal vaccination. Further follow-up is needed to assess late-onset outcomes and the effect of later circulating SARS-CoV-2 variants. Continued investment in linked maternal-child health data is essential to monitor both immediate and long-term effect of emerging infections during pregnancy.

**Summary box:** *What is already known on this topic:* Existing research has shown a higher risk of preterm births in children who were exposed to SARS-CoV-2 in utero, but health outcomes beyond the neonatal period are not well described. Published evidence on comparing the effects of exposure to different variants in utero on children’s health outcomes remains limited.

*What this study adds:* This study found no evidence of an increased hazard of first emergency or planned hospital admissions up to 40 months of age associated with in utero exposure to SARS-CoV-2 during the wild-type or Alpha periods, in the absence of maternal vaccination.

## Introduction

SARS-CoV-2-related disease has been shown to be more severe in pregnant than non-pregnant women[1]. However, much less is known about the outcomes for children who were exposed to SARS-CoV-2 during pregnancy. According to the Developmental Origins of Health and Disease hypothesis, periconceptual and foetal exposures may cause irreversible genetic and epigenetic programming changes, increasing the risk of chronic diseases later in life[2,3]. Maternal respiratory viral infections during pregnancy have been shown to be associated with a higher risk of neurodevelopmental disorders[4,5] and leukaemia in offspring[6–8], although findings are not always consistent across studies. Vertical transmission is one mechanism by which maternal infection in pregnancy affects foetus and child, but this is rare in the case of SARS-CoV-2[9–12]. However, maternal infections in pregnancy, including SARS-CoV-2, even in the absence of vertical transmission, may also negatively affect foetal and infant outcomes through pathways linked to immune activation and inflammation, including inflammatory cytokines signalling, placental dysfunction, and altered foetal immune programming[13–15]. In a meta-analysis of 117 studies, an estimated 7% (95% confidence interval: 6% to 8%) of pregnant women globally were infected with SARS-CoV-2 between December 2019 and April 2021[1]. Given the global scale of the pandemic and the continued circulation of SARS-CoV-2 virus, the potential long-term consequences for child health warrant rigorous investigation.

A systematic review including 204 studies from 36 countries, mostly only including children exposed in utero to SARS-CoV-2 in the third trimester, demonstrated inconsistent findings with respect to adverse birth and neonatal outcomes. However, the review reported a pooled estimate of 1.93 (95% confidence interval [CI]: 1.33 to 2.80) times higher odds of premature delivery and 3.35 (0.84 to 13.36) times higher odds of neonatal intensive care unit (NICU) admission among children exposed to SARS-CoV-2 in utero[16]. The authors attributed the increased NICU admission rate to prematurity, for isolation and observation purposes, or for care of a baby whose mother could not care for the baby herself due to COVID-19 disease[16]. Further studies, mostly with limited sample size, suggest that infants whose mothers had SARS-CoV-2 infection in pregnancy may have an increased risk of respiratory disease in the neonatal period and developmental disorders in infancy, including developmental disorders of speech and language and motor function[16–18]. No association has been shown between SARS-CoV-2 infection during the first trimester of pregnancy and risk of congenital anomalies in children[19]. The vast majority of existing studies have focused on health outcomes in the perinatal and neonatal period[16,17,20–23]. While a few studies from high-income countries have followed children beyond this period, they have typically examined a limited set of prespecified outcomes, mostly neurological disorders[18,19,24–27].

We aimed to examine the impact of in utero exposure to maternal SARS-CoV-2 infection during pregnancy on children’s emergency and planned hospital admissions up to age 40 months in England. We focused on women infected during the wild-type and Alpha waves of the COVID-19 pandemic, before the recommendation that pregnant women receive COVID-19 vaccines[28,29].

## Data and Methods

### Data sources

This cohort study used multiple linked national administrative health datasets, including Hospital Episode Statistics Admitted Patient Care dataset (HES APC, from here on referred to as HES for brevity) for hospital admissions, Second Generation Surveillance System (SGSS, ‘Pillar 1’l; see below for an explanation of the pillars) and Non-hospital Antigen Testing Results (‘Pillar 2’) for SARS-CoV-2 testing, the birth notification, death registration, and mother-baby linkage developed by Feng et al.[30,31]. Figure 1 illustrates the linkage between these datasets and how we derived exposure and outcome variables. A description of data sources and approaches to identify birth cohorts and estimate date of birth is presented in Supplementary Methods (Table A1 and Figure A1).

**Figure 1.**
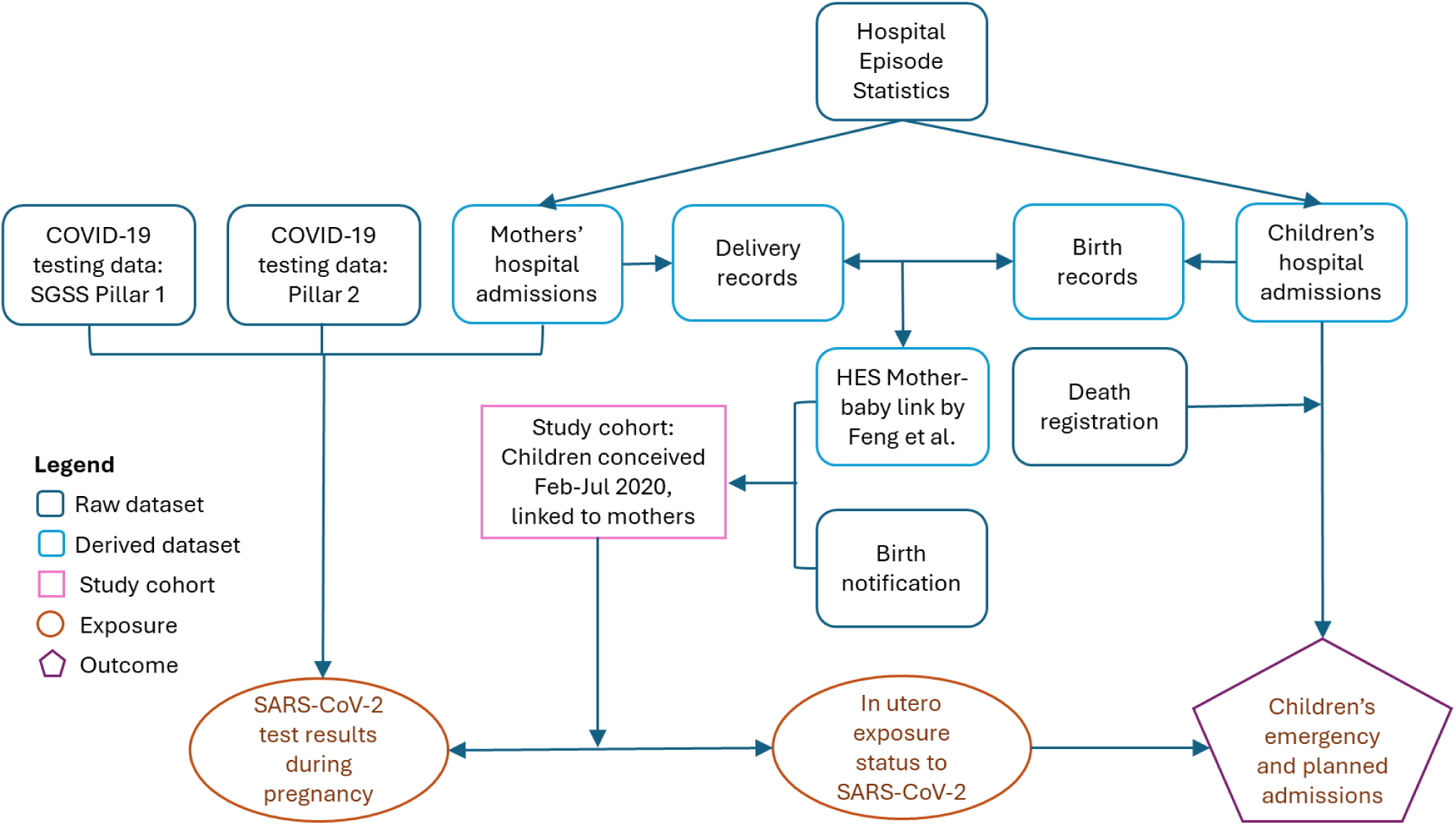
Linkage between data sources to derive the exposure and outcome variables.

We derived a national birth cohort of all babies born in England from birth notifications[32] and a HES mother-baby linked dataset within HES[30,31]. Births recorded in either source were included. Children were followed up using HES data and death registration data linked to the birth cohort. Cleaning and linking HES episodes into continuous inpatient admissions episodes used an algorithm developed by Hardelid et al [33,34] (Table A2 in Supplementary Methods). Hospital transfers and admissions within one day were treated as one inpatient admission.

Maternal SARS-CoV-2 infections were determined via linkage between maternal HES records and Pillar 1 and 2 datasets that contain information on SARS-CoV-2 test results. Pillar 1 was introduced in March 2020, initially prioritising individuals with a medical need and critical key workers. This dataset was expanded in May 2020 following the introduction of universal testing of all hospital-admitted patients, including admissions to maternity services for delivery. This routine asymptomatic testing for hospital-admitted patients came to an end in August 2022[35,36]. The Pillar 1 dataset contains data on positive SARS-CoV-2 tests reported by NHS virology laboratories to the Public Health England (the national infectious disease surveillance agency), predominantly based on polymerase chain reaction (PCR) testing[37–39].

Pillar 2 community testing was launched in April 2020, initially using PCR testing for symptomatic key workers and general public via local test centres[40]. Lateral flow testing (rapid antigen tests) was introduced in December 2020 and gradually expanded to include asymptomatic individuals, with universal access implemented in April 2021 continuing until April 2022[41–43]. The Pillar 2 dataset contains data on positive and negative SARS-CoV-2 tests conducted in the community (e.g., home testing, drive-through and mobile test centres) [37–39,44]. Note that lateral flow test results conducted at home relied on individuals’ self-reporting test results online. Key changes to Pillar 1 and Pillar 2 testing policies were summarised in Figure 2, and the individuals included in each exposure group during the intervals between policy changes are described in Table B1 in Supplementary Tables and Figures.

**Figure 2.**
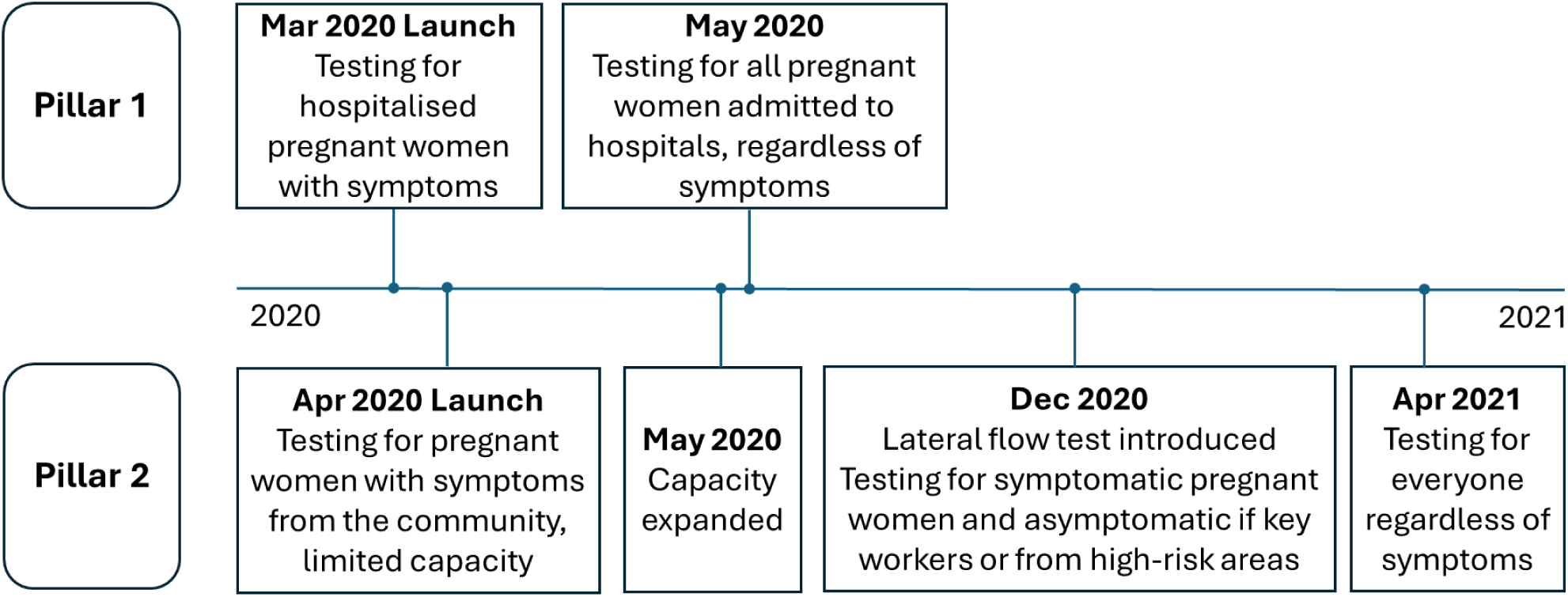
Key changes in Pillar 1 and Pillar 2 testing policy relevant to the exposure period.

### Study population

We included all babies in the birth cohort with an estimated conception date between 1^st^ February and 31^st^ July 2020, ensuring most births occurred before the COVID-19 vaccine rollout. The date of conception was defined as 2+0 weeks gestation[45], where the start of gestation was calculated using the estimated date of birth and gestational age at birth in weeks. We defined the birth cohort by date of conception rather than date of birth to avoid unbalanced distribution on gestational age and thus reduce fixed cohort bias[46]. Multiple births, stillbirths, and children whose mothers were residents outside England at delivery were excluded. Rules for identifying stillbirths and multiple births in HES are presented in Table A3 in Supplementary Methods.

### Outcomes

The primary outcome was children’s first emergency admission after discharge from delivery. We had two secondary outcomes: children’s first planned (elective) admission and overall admission rates (including emergency and planned) according to primary diagnosis. Emergency and planned admissions were distinguished using the ‘admimeth’ variable (Table A4 in Supplementary Methods). Birth admissions, between-hospital transfer, and unclassified admissions were excluded from follow-up.

### Exposure

The primary exposure was exposure to SARS-CoV-2 variants in utero, determined by combining the mother’s information from Pillar 1, Pillar 2 and hospital diagnoses in HES during pregnancy. SARS-CoV-2 infections identified through linked testing were supplemented by COVID-19 diagnoses recorded in maternal HES records (International Classification of Diseases 10^th^ version, ICD-10 codes U07.1-U07.2) during pregnancy, with the episode start date considered as the date of infection. De-duplication and cleaning of Pillar 1 and 2 data are described in Supplementary Methods (Table A5 and A6).

The first positive test or COVID-19 diagnosis since conception was defined as the start of the first SARS-CoV-2 infection during pregnancy. Two positive tests 90 days or less apart were considered part of the same infection episode[47]. Children whose mothers were tested for SARS-CoV-2 during pregnancy with unknown results were excluded from the analysis. Children exposed more than once in utero in both the wild-type and the Alpha-period were excluded.

In utero positive exposure status was classified according to the dominant circulating variant at the time of the first positive test or the first admission date with an ICD-10 diagnosis of SARS-CoV-2 infection within each infection episode during pregnancy: “positive, wild-type” (from outbreak start to 15^th^ December 2020) or “positive, Alpha” (16^th^ December 2020 to 15^th^ May 2021[48–50]). Children whose mothers had a negative test result in Pillar 2 at any point during pregnancy and no positive test result or diagnosis throughout pregnancy were classified as the “test-negative” group, irrespective of timing of the negative test; and the remaining children whose mothers had no record of either a positive or negative test result, nor a diagnosis in HES during pregnancy were classified as the “no recorded result” group. The four exposure groups are mutually exclusive. All pregnancies included in this study had progressed beyond the first trimester before the Alpha period began, due to the conception date inclusion criterion.

### Follow up

Children were followed from the day after their estimated date of birth until the first occurrence of the outcome event, death, or reaching 40 months of age, whichever occurred first. To estimate admission rates, we included admissions from the day after their estimated date of birth until death or reaching 40 months of age, whichever occurred first.

### Covariates

We used Directed Acyclic Graphs to identify confounders for inclusion in the statistical models (Figure A2 in Supplementary Methods)[51]. Gestational age (preterm birth: <37 weeks, term birth: 37-42 weeks, post-term birth: >42 weeks) lies on the causal pathway between in utero exposure to SARS-CoV-2 and child hospital admission, so it was not considered a confounder. However, we present the distribution of gestational ages in each exposure group. To minimise missing data for these covariates, we integrated information from children’s HES admissions records within the first month of life, birth notification, mother’s delivery records, and the mother-baby linkage[31] when available. The included confounders, definitions and the source of information are presented in Table 1.

**Table 1.** Definition of confounders and the source of information.

| Confounders | Definition | Source of information |
| --- | --- | --- |
| Mother’s ethnicity | Ethnicity of the mother: White, Asian, Black, mixed, or other[52] | Mothers’ HES delivery records; if missing, replace with children’s ethnicity in children’s HES admissions within 28 days of birth; if missing, replace with children’s ethnicity in birth notification. |
| Area deprivation decile | Deciles of the Index of Multiple Deprivation (IMD), measured at the Lower Super Output Area (LSOA) level (an average size of 1500 residents or 650 households[52–54]) | Area deprivation decile in mother’s HES delivery records; if missing, replace with area deprivation decile in children’s HES admissions within 28 days of birth; if missing, replace with IMD decile in the mother-baby link developed by Feng et al. |
| Maternal chronic condition | Presence of chronic condition defined by Charlson Comorbidities Classification based on diagnoses recorded in HES up to three years before conception[55,56] | ICD-10 disease codes in mother's HES admissions up to 3 years before conception. |
| Maternal age | Age of the mother at delivery: <25, 25-29, 30-34, 35-39, and $\geq 40$ years | The most common value in children's HES admissions within 28 days of birth; if missing, replace with maternal age from the mother-baby link developed by Feng et al.; if missing, replace with maternal age in the mother's HES delivery admission. |
| Whether the child was a London resident | Government office region, LSOA, or postcode district is Greater London area. | The most common value in children's admissions within 28 days of birth; if missing, replace with mother's residence area in delivery records. |
| Month and year of conception | Month-year of the estimated date of conception | Children's birth admissions in HES and birth notification. |

## Statistical analysis

We plotted the number of conceptions, births, in utero exposure to SARS-CoV-2, and emergency and planned admissions against calendar dates. To understand testing policies over time, we also plotted the number of SARS-CoV-2 tests and COVID-19 diagnoses recorded for mothers in each dataset over the study period. We compared the distribution of the key variables of interest and the extent of missing data among children in the birth cohort based on their in utero SARS-CoV-2 exposure status (positive, wild-type; positive, Alpha; test-negative; no recorded result). We plotted the Kaplan-Meier curves for time to first emergency and planned admission. We calculated rates of emergency admissions and planned admissions per 1000 child years, respectively, stratified by exposure status (including all admissions, not just the first; details in Supplementary Methods).

We examined the association between in utero SARS-CoV-2 exposure and time to first emergency or planned admission using Cox proportional hazards regression models with robust standard errors, separately for emergency and planned admissions. For each outcome, we first fitted an unadjusted model including the main exposure (exposure to SARS-CoV-2; coded into four groups: positive, wild-type; positive, Alpha; test-negative; no recorded result), followed by an adjusted model incorporating potential confounders listed in Table 1. The proportional hazards assumption was assessed by comparing models with and without an interaction term between exposure status and time (child’s age), using likelihood ratio tests.

We also conducted trimester-specific analyses among children with a record of in utero exposure to SARS-CoV-2. Trimester of exposure was determined by the timing of the first positive test result of each episode of infection. We constructed Cox proportional hazard regression models among children exposed in utero to examine the effect of timing of in utero exposure on the time to first emergency or planned admissions. Models initially included the variant period of exposure (wild-type vs. Alpha period) and the trimester of exposure (first trimester: conception to 12 weeks; second trimester [reference]: 13 to 27 weeks; third trimester: 28 weeks to birth), and were subsequently adjusted for confounders listed in Table 1. Children who were unexposed or whose mothers had multiple episodes of infection in different trimester were excluded from the trimester-specific analyses.

To calculate admission rates by primary diagnosis, we classified the primary diagnosis of the first episode of each admission into disease groups according to ICD-10 chapters[57] (Table A7 in Supplementary Methods). Admission rates were calculated for each disease group, including all emergency and planned admissions per 1000 child years.

To account for multiple tests, we used 99% confidence intervals (CI). All counts smaller than five were reported as “<5” to minimise the potential of re-identification[58]. All regression models included only children with complete data for all covariates. All analyses were done using STATA 18.0 (StataCorp LLC, College Station, Texas). The Strengthening the Reporting of Observational Studies in Epidemiology (STROBE) checklist is presented in Supplementary STROBE Checklist)[59].

## Sensitivity analysis

To rule out the influence of hospital admissions due to COVID-19, we performed a sensitivity analysis excluding COVID-19-related admissions in children (definitions provided in Supplementary Sensitivity Analysis). To address the potential violation of the proportional hazard assumption, we performed a sensitivity analysis using logistic regression models, assuming equal follow-up time for all children. To account for potential overestimation of person-time caused by excluding neonatal admissions immediately after birth, we performed a sensitivity analysis excluding admissions within the first 7 days of life, with follow-up starting from day 8. To address potential selection bias arising from changes in SARS-CoV-2 testing policies and capacities across variant periods, models were constructed and admissions rates calculated separately for the wildtype and Alpha periods, using children whose mothers tested negative during pregnancy within the corresponding variant period as the comparator group.

## Patient and public involvement

Whilst patients and the public have not been involved with this specific piece of research, the study team met with the Great Ormond Street Hospital Biomedical Research Centre Parents’ and Carers’ Advisory Group in April 2022 to discuss the overall project and discuss the use of linked administrative health data to address research questions relating to the child and maternal health impacts of SARS-CoV-2 infection.

## Results

This study included 262,086 children born between June 2020 and May 2021 to 261,797 mothers (Figure 3 and top panel of Figure 4). Table 2 summarises children’s characteristics by exposure status. A higher proportion of children exposed in utero were from ethnic minority groups, compared to children in the no recorded result and test-negative groups. Children exposed in utero were more likely to be born in deprived areas than children in the no-recorded-result and test-negative groups (percentage of children from the 10% most deprived areas: 19.7% in the wild-type group and 16.8% in the Alpha group vs. 13.8% in the no-recorded-result group and 11.4% in the test-negative group). The test-negative group had a higher proportion of children whose mothers had chronic conditions (4.8%) and tended to be conceived later than the other three groups.

**Figure 3.**
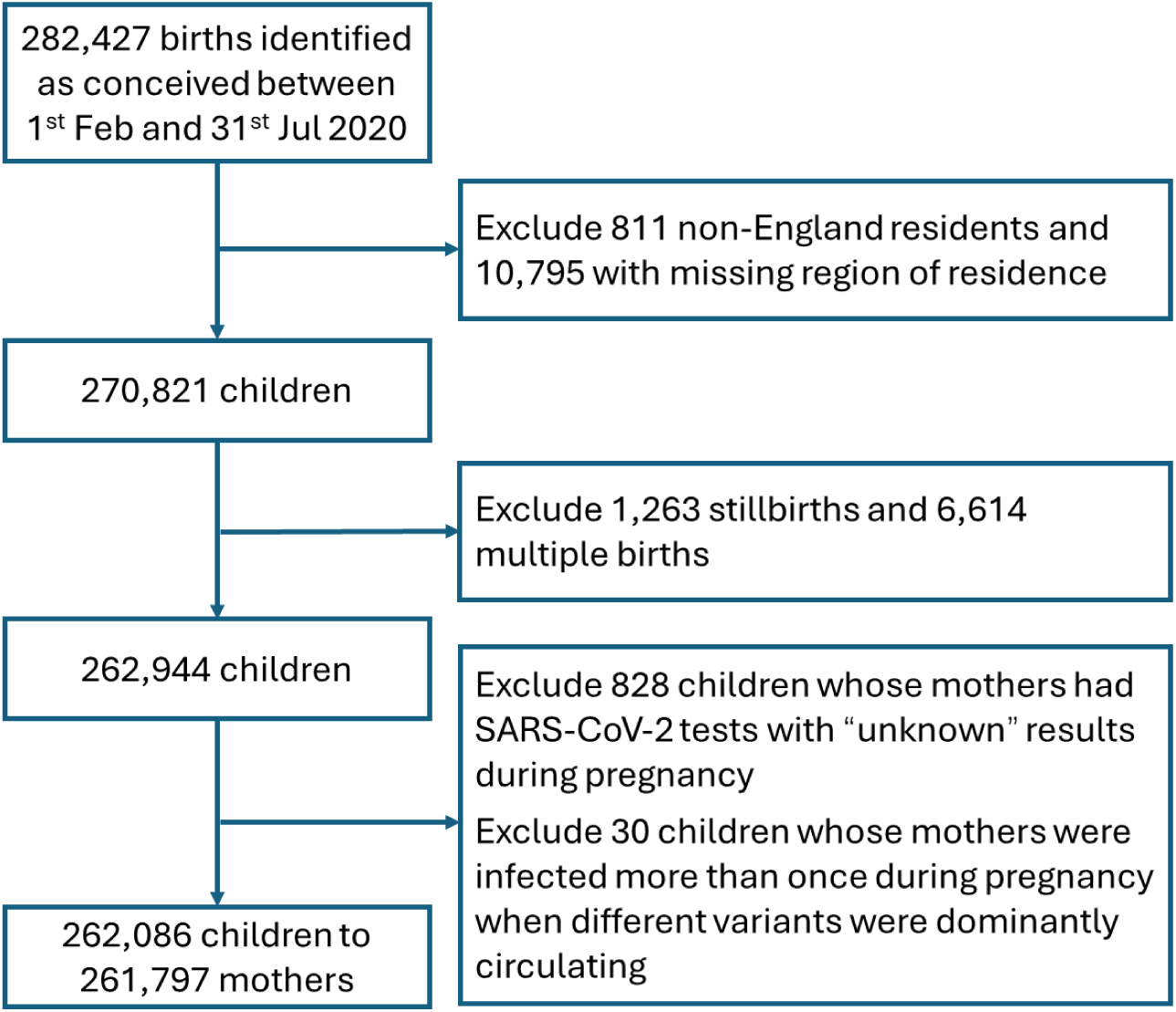
Flow chart showing the inclusion of births in the birth cohort.

**Figure 4.**
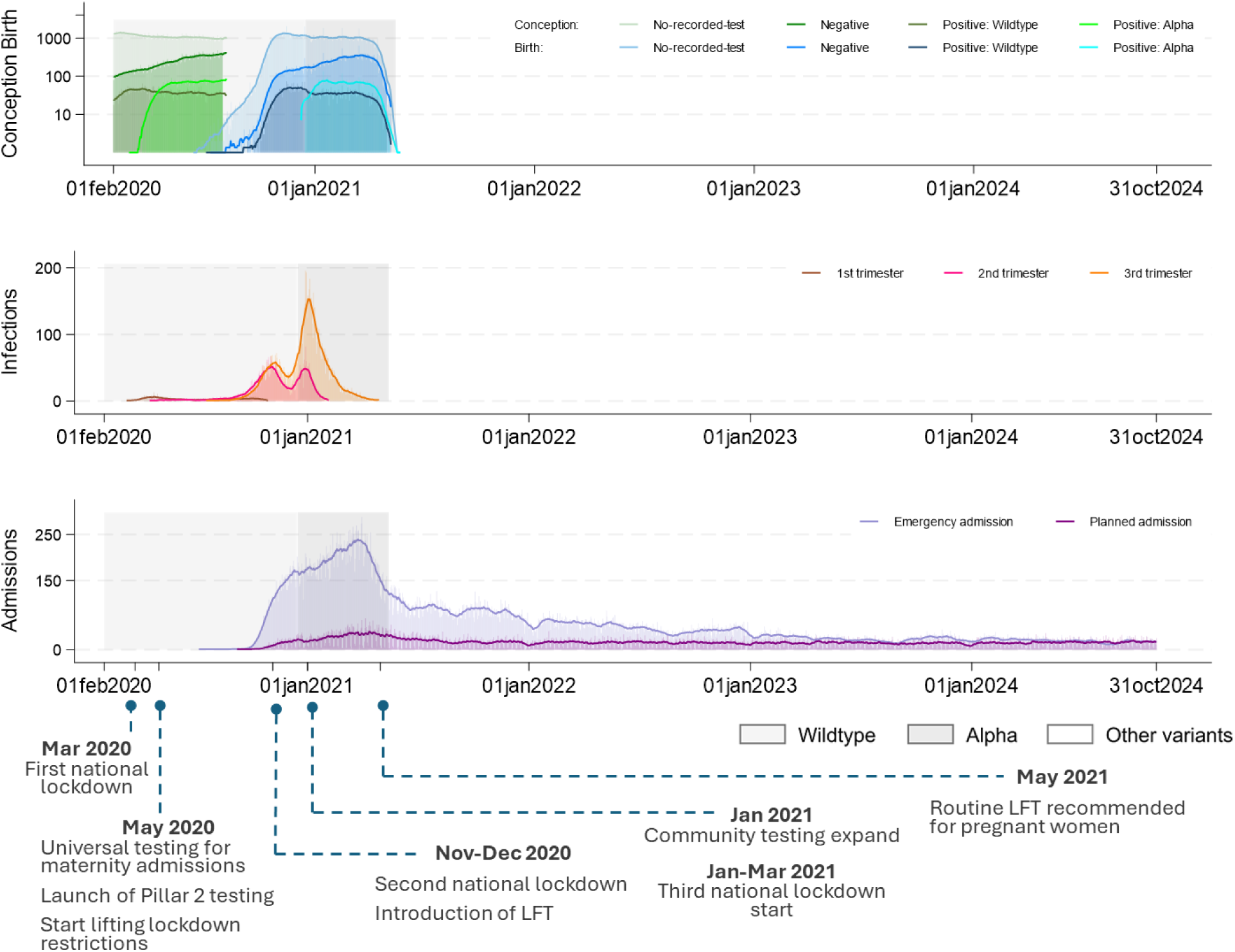
Distribution of conceptions, births, maternal SARS-CoV-2 infections in pregnancy and emergency and planned admissions in the study cohort. From top panel to bottom panel: the number of conceptions and births; the number of maternal SARS-CoV-2 infections in pregnancy; the number of emergency admissions and planned admissions in children. Transparent bars show daily counts, and solid lines show the 15-day rolling average of daily counts (a week before + the index day + a week after).

**Table 2.** Distribution of the characteristics by in-utero exposure status to SARS-CoV-2 during pregnancy.

| Characteristics | Number of children (column percentage) |  |  |  | Total (column percentage) |
| --- | --- | --- | --- | --- | --- |
|  | Test-negative | Positive, wild-type | Positive, Alpha | No recorded result |  |
| Number of children | 43,225 | 6,888 | 7,829 | 204,144 | 262,086 |
| Number of children with emergency admission | 16,023 (37.1) | 2,646 (38.4) | 2,634 (33.6) | 67,886 (33.3) | 89,189 (34.0) |
| Number of children with planned admission | 4,164 (9.6) | 716 (10.4) | 745 (9.5) | 19,010 (9.3) | 24,635 (9.4) |
| **Area of residence** |  |  |  |  |  |
| London | 6,294 (14.6) | 983 (14.3) | 2,266 (28.9) | 38,979 (19.1) | 48,522 (18.5) |
| Non-London | 36,931 (85.4) | 5,905 (85.7) | 5,563 (71.1) | 165,165 (80.9) | 213,564 (81.5) |
| **Ethnicity of mother** |  |  |  |  |  |
| White | 35,647 (82.5) | 4,572 (66.4) | 5,151 (65.8) | 151,853 (74.4) | 197,223 (75.3) |
| Asian | 4,269 (9.9) | 1,533 (22.3) | 1,514 (19.3) | 27,189 (13.3) | 34,505 (13.2) |
| Black | 1,125 (2.6) | 278 (4.0) | 530 (6.8) | 10,552 (5.2) | 12,485 (4.8) |
| Mixed | 1,011 (2.3) | 175 (2.5) | 210 (2.7) | 5,443 (2.7) | 6,839 (2.6) |
| Others | 1,172 (2.7) | 330 (4.8) | 424 (5.4) | 9,100 (4.5) | 11,026 (4.2) |
| Missing | <5 (<0.05) | 0 (0) | 0 (0) | 7 (<0.05) | 8 (<0.01) |
| **Area deprivation decile** |  |  |  |  |  |
| Most deprived 10% | 4,923 (11.4) | 1,359 (19.7) | 1,315 (16.8) | 28,177 (13.8) | 35,774 (13.7) |
| 10% to 20% | 4,788 (11.1) | 975 (14.2) | 1,213 (15.5) | 25,775 (12.6) | 32,751 (12.5) |
| 20% to 30% | 4,650 (10.8) | 820 (11.9) | 1,003 (12.8) | 23,700 (11.6) | 30,173 (11.5) |
| 30% to 40% | 4,466 (10.3) | 732 (10.6) | 886 (11.3) | 21,489 (10.5) | 27,573 (10.5) |
| More deprived 40% to 50% | 4,273 (9.9) | 614 (8.9) | 720 (9.2) | 19,742 (9.7) | 25,349 (9.7) |
| Less deprived 40%-50% | 4,264 (9.9) | 572 (8.3) | 659 (8.4) | 18,254 (8.9) | 23,749 (9.1) |
| 30%-40% | 4,068 (9.4) | 512 (7.4) | 547 (7.0) | 17,269 (8.5) | 22,396 (8.6) |
| 20% to 30% | 4,002 (9.3) | 486 (7.1) | 560 (7.2) | 16,935 (8.3) | 21,983 (8.4) |
| 10% to 20% | 4,026 (9.3) | 442 (6.4) | 486 (6.2) | 16,977 (8.3) | 21,931 (8.4) |
| Least deprived 10% | 3,758 (8.7) | 375 (5.4) | 437 (5.6) | 15,800 (7.7) | 20,370 (7.8) |
| Missing | 7 (0.02) | <5 (<0.05) | <5 (<0.05) | 26 (0.01) | 37 (0.01) |
| **Mother's chronic condition** |  |  |  |  |  |
| Yes | 2,063 (4.8) | 264 (3.8) | 267 (3.4) | 7,175 (3.5) | 9,769 (3.7) |
| No | 41,162 (95.2) | 6,624 (96.2) | 7,562 (96.6) | 196,969 (96.5) | 252,317 (96.3) |
| **Year-Month of conception** |  |  |  |  |  |
| Feb 2020 | 3,307 (7.7) | 1,011 (14.7) | 0 (0) | 39,098 (19.2) | 43,416 (16.6) |
| Mar 2020 | 4,564 (10.6) | 1,403 (20.4) | 98 (1.3) | 37,195 (18.2) | 43,260 (16.5) |
| Apr 2020 | 5,844 (13.5) | 1,163 (16.9) | 1,163 (14.8) | 32,654 (16.0) | 40,824 (15.6) |
| May 2020 | 8,135 (18.8) | 1,117 (16.2) | 2,099 (26.8) | 32,549 (15.9) | 43,900 (16.8) |
| Jun 2020 | 10,150 (23.5) | 1,147 (16.7) | 2,164 (27.6) | 32,006 (15.7) | 45,467 (17.4) |
| Jul 2020 | 11,225 (26.0) | 1,047 (15.2) | 2,305 (29.4) | 30,642 (15.0) | 45,219 (17.3) |
| **Maternal age** |  |  |  |  |  |
| <25 | 5,775 (13.4) | 1,090 (15.8) | 1,382 (17.7) | 30,868 (15.1) | 39,115 (14.9) |
| 25-29 | 11,445 (26.5) | 2,039 (29.6) | 2,130 (27.2) | 53,271 (26.1) | 68,885 (26.3) |
| 30-34 | 15,380 (35.6) | 2,208 (32.1) | 2,529 (32.3) | 68,275 (33.4) | 88,392 (33.7) |
| 35-39 | 8,079 (18.7) | 1,142 (16.6) | 1,318 (16.8) | 38,741 (19.0) | 49,280 (18.8) |
| >=40 | 1,772 (4.1) | 298 (4.3) | 328 (4.2) | 9,204 (4.5) | 11,602 (4.4) |
| N missing values | 774 (1.8) | 111 (1.6) | 141 (1.8) | 3,785 (1.9) | 4,812 (1.8) |
| **Gestational age (weeks)** |  |  |  |  |  |
| Preterm birth (<37) | 2,071 (4.8) | 460 (6.7) | 615 (7.9) | 12,998 (6.4) | 16,144 (6.2) |
| Term birth (34-42) | 41,126 (95.1) | 6,421 (93.2) | 7,205 (92.0) | 191,007 (93.6) | 245,759 (93.8) |
| Post-term birth (>42) | 28 (0.1) | 7 (0.1) | 9 (0.1) | 139 (0.1) | 183 (0.1) |

The number of positive tests increased markedly at the onset of the Alpha wave in January 2021, coinciding with the expansion of community testing and wider use of Pillar 2 testing (Figure B1 in Supplementary Tables and Figures).

A total of 43,225 (16.5%) children were in the test-negative exposure group, while 5.6% were exposed to SARS-CoV-2 in utero at least once, with 6,888 (2.6%) exposed in the wild-type period and 7,829 (3.0%) in the Alpha period. Among exposed children, the positive maternal test was reported in the second trimester for 29.5% (n=4,343), in the third for 67.4% (n=9,925), with only 4.1% of children exposed in the first trimester (Figure 4). Ten mothers tested positive in both the first and second trimesters, <5 in the first and the third trimesters, and 140 in the second and third trimesters.

In total, 89,189 children (34.0%) had at least one emergency admission before age 40 months: 55,663 (21.2%) had a single emergency admission, while 33,526 (12.8%) had two or more. A total of 24,635 children (9.4%) had at least one planned admission, with 16,881 (6.4%) having one planned admission and 7,754 (3.0%) having two or more (Table 2 and Figure 4). About two-thirds (n=162,293; 61.9%) of children had no admissions, while 14,031 (5.4%) children had both emergency and planned admissions. More than half (n=14,031; 57.0%) of children with a planned admission also had an emergency admission. Kaplan-Meier curves for emergency and planned admissions are shown in Figures B2 and B3 in the Supplementary Tables and Figures.

Children with in utero exposure during the Alpha period and those without a maternal test result had lower emergency admission rates (169.6 per 1000 child-years [99% CI: 163.2 to 176.3] and 170.7 [169.4 to 172.0], respectively) than those exposed during the wild-type period and those unexposed during either period (209.1 [201.4 to 217.0] and 200.4 [197.4 to 203.5] respectively). Planned admission rates were lower among children exposed in utero during the Alpha period (45.4 [42.1 to 48.9]), while rates were similar across the other three groups (Table 3).

**Table 3.** Hospital admission rates by in utero SARS-CoV-2 exposure status (all admissions other than birth admissions were included)

| In-utero exposure status | Number of children | Number of child-year (*1000) | Number of admissions | Rate per 1000 child-years (99% CI) |
| --- | --- | --- | --- | --- |
| **Emergency admissions** |  |  |  |  |
| Test-negative | 43,225 | 143.8 | 28,825 | 200.4 (197.4, 203.5) |
| Positive, Wild-type | 6,888 | 22.9 | 4,786 | 209.1 (201.4, 217.0) |
| Positive, Alpha | 7,829 | 26.1 | 4,418 | 169.6(163.2, 176.3) |
| No-recorded-result | 204,144 | 678.4 | 115,783 | 170.7 (169.4, 172.0) |
| Total | 262,086 | 871.2 | 153,812 | 176.6 (175.4, 177.7) |
| **Planned admission** |  |  |  |  |
| Test-negative | 43,225 | 143.9 | 7,346 | 51.1 (49.5, 52.6) |
| Positive: Wild-type | 6,888 | 22.9 | 1,258 | 54.9 (51.1, 59.1) |
| Positive: Alpha | 7,829 | 26.1 | 1,183 | 45.4 (42.1, 48.9) |
| No-recorded-result | 204,144 | 678.6 | 33,979 | 50.1 (49.4, 50.8) |
| Total | 262,086 | 871.5 | 43,766 | 50.2 (49.6, 50.8) |

A total of 257,241 children (98.2%) with complete data for all covariates were included in the Cox proportional hazard regression models. There was no evidence for a difference in the time to the first emergency admission between children exposed in utero during the wild-type period and children unexposed in either period (i.e., test-negative group) (Figure 5 and Table B2 in Supplementary Tables and Figures). However, children exposed during the Alpha variant period and those in the no-recorded-result group had 8% (adjusted HR=0.92, 99% CI: 0.87 to 0.97) and 12% (aHR=0.88, 0.86 to 0.90) lower hazard of first emergency admission than the test-negative group, respectively.

**Figure 5.**
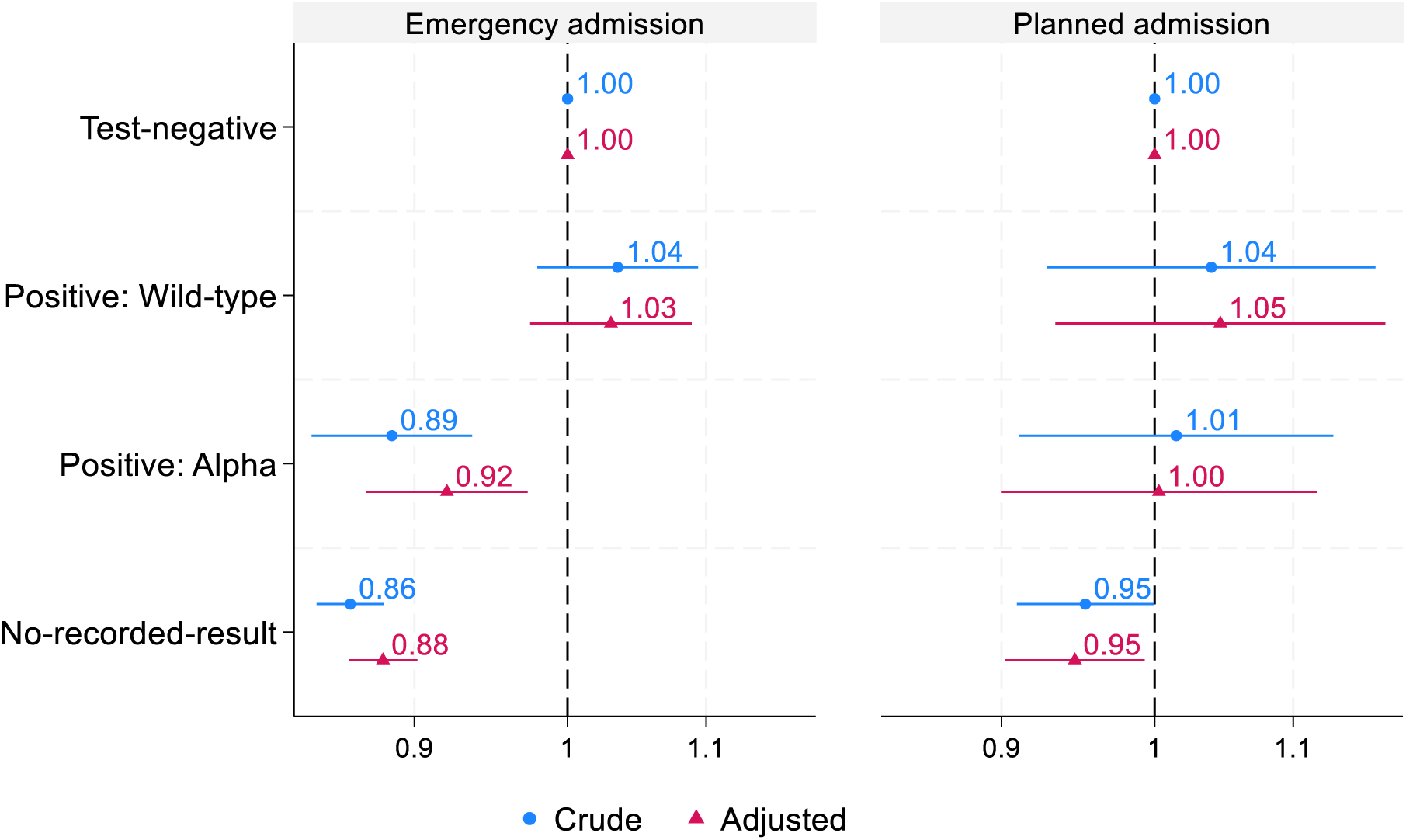
Time to child’s first emergency and planned admission up to 40 months old: hazard ratio and 99% confidence interval by in utero SARS-CoV-2 exposure status, children conceived and born between February 2020 and April 2021 in England. Reference=Test-negative group. Note: Results based on Cox proportional hazard regression models. Adjusted models include maternal ethnicity, area deprivation decile, mother’s history of chronic conditions, maternal age, London residence, and month of conception. A total of 257,249 children with complete data for all covariates were included.

Children in the no-recorded-result group had a 5% lower hazard of first planned admission compared with the test-negative group, while no significant difference was observed between the two positive groups and the test-negative group, regardless of adjustment (Figure 5 and Table B2). The proportional hazard assumption was met for the model examining emergency admissions but was violated for the planned admissions model in the no-recorded-result group (Table B3).

Only 603 children were exposed during the first trimester, all during the wild-type period. Similar numbers of exposed children were observed during the wild-type period in the second (n=3,035) and third trimesters (n=3,333) (Table 4). A total of 14,306 children with confirmed in utero exposure and complete data for all covariates were included in the trimester-specific regression analyses, after excluding 154 children where exposure spanned two trimesters.

**Table 4.** Emergency and planned admissions by trimester of in utero exposure to SARS-CoV-2.

| Trimester of in utero exposure | Positive: Wild-type |  | Positive: Alpha |  |
| --- | --- | --- | --- | --- |
|  | N children | N children with a hospital admission (%) | N children<sup>#</sup> | N children with a hospital admission (%) |
| **Emergency admission** |  |  |  |  |
| First trimester | 603 | 213 (35.3) | - | - |
| Second trimester | 3,035 | 1,066 (35.1) | 1,308 | 396 (30.3) |
| Third trimester | 3,333 | 1,124 (33.7) | 6,592 | 1,973 (29.9) |
| **Planned admission** |  |  |  |  |
| First trimester | 603 | 47 (7.8) | - | - |
| Second trimester | 3,035 | 206 (6.8) | 1,308 | 96 (7.3) |
| Third trimester | 3,333 | 232 (7.0) | 6,592 | 442 (6.7) |

<sup>#</sup> As this study includes children conceived between Feb and Jul 2020 and the Alpha period starts in Dec 2020, no child in this study would have in utero exposure in the first trimester during the Alpha period.

Among in utero exposed children, no evidence was found for a difference in time to first emergency or planned admissions by trimester of exposure, while children in utero exposed during the Alpha period showed a 16% lower hazard of the first emergency admission, compared with those exposed during the wild-type period (aHR=0.84, 0.75 to 0.93) (Figure 6).

**Figure 6.**
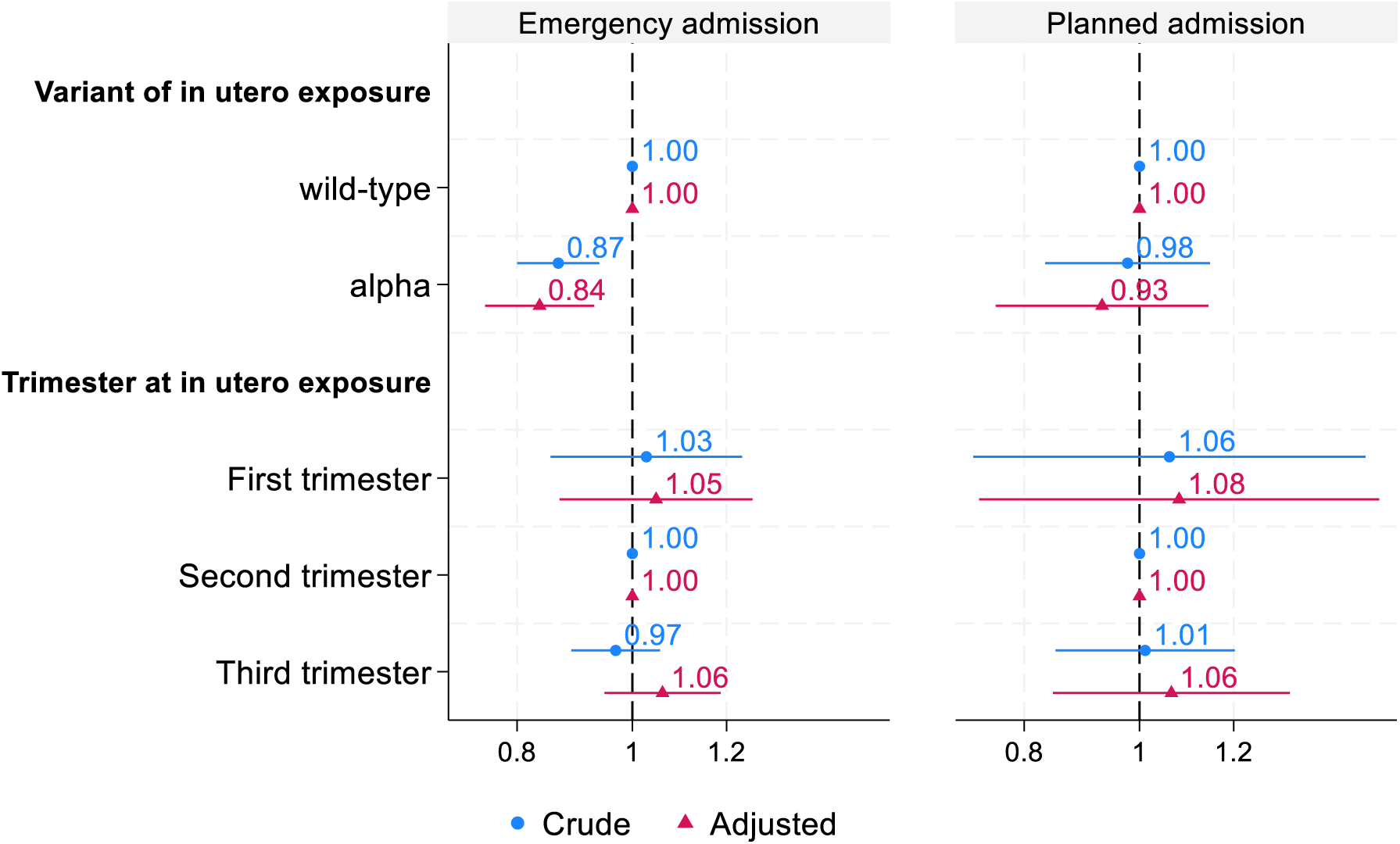
The impact of variant and timing of SARS-CoV-2 exposure in utero on time to the first emergency and planned admissions. Note: Results based on Cox proportional hazard regression models. Adjusted models include maternal ethnicity, deprivation decile, maternal chronic conditions, maternal age, London residence, and month of conception. A total of 14,306 children with complete data for all covariates were used.

Children with in utero exposure during the wild-type period had the highest admission rate (264.1 per 1000 child-years, 99% CI: 255.5 to 272.9), followed by those whose mothers tested negative during pregnancy (251.5, 248.2 to 255.0). Lower admission rates were observed among children exposed in utero during the Alpha period (215.0, 207.7 to 222.5) and those with no recorded maternal test during pregnancy (220.8, 219.3 to 222.2) (Table 5).

**Table 5.** Admission rates per 1000 child-years (99% CI) by primary diagnosis, including all emergency and planned admissions up to 40 months old.

| Disease group | Number of disease cases | Number of admissions | Test-negative | Positive: Wild-type | Positive: Alpha | No-recorded-result | Total |
| --- | --- | --- | --- | --- | --- | --- | --- |
| **Certain infectious and parasitic diseases** |  |  |  |  |  |  |  |
| Bacterial infectious diseases | 7,692 | 8,880 | 11.6 (10.9,12.4) | 14.0 (12.1,16.2) | 9.9 (8.4,11.6) | 9.8 (9.5,10.1) | 10.2 (9.9,10.5) |
| Non-bacterial infectious diseases | 13,955 | 19,337 | 24.0 (23.0,25.1) | 27.7 (25.0,30.7) | 20.6 (18.4,23.0) | 21.7 (21.2,22.1) | 22.2 (21.8,22.6) |
| **Neoplasms** |  |  |  |  |  |  |  |
| Malignant neoplasms | 193 | 4,420 | 5.3 (4.8,5.8) | 11.0 (9.3,12.9) | -# | 5.0 (4.8,5.2) | 5.1 (4.9,5.3) |
| Benign neoplasms | 906 | 1,501 | 2.5 (2.2,2.8) | 1.1 (0.7,1.8) | 1.9 (1.3,2.8) | 1.6 (1.5,1.7) | 1.7 (1.6,1.8) |
| **Diseases of the blood and blood-forming organs and certain disorders involving the immune mechanism** | 806 | 2,502 | 1.9 (1.6,2.2) | 4.4 (3.4,5.7) | 2.8 (2.0,3.7) | 3.0 (2.9,3.2) | 2.9 (2.7,3.0) |
| **Endocrine, nutritional and metabolic diseases** | 1,228 | 2,373 | 3.0 (2.7,3.4) | 2.2 (1.5,3.1) | 2.6 (1.9,3.6) | 2.7 (2.5,2.8) | 2.7 (2.6,2.9) |
| **Mental and behavioural disorders** | 330 | 360 | 0.4 (0.3,0.6) | 0.2 (0.0,0.6) | 0.3 (0.1,0.7) | 0.4 (0.4,0.5) | 0.4 (0.4,0.5) |
| **Diseases of the nervous system** | 1,902 | 3,435 | 4.1 (3.7,4.6) | 5.0 (4.0,6.4) | 3.8 (3.0,5.0) | 3.9 (3.7,4.1) | 3.9 (3.8,4.1) |
| **Diseases of the eye and adnexa** | 1,596 | 1,907 | 2.4 (2.1,2.7) | 2.6 (1.9,3.7) | 1.9 (1.3,2.8) | 2.1 (2.0,2.3) | 2.2 (2.1,2.3) |
| **Diseases of the ear and mastoid process** | 2,258 | 2,626 | 3.6 (3.2,4.0) | 3.9 (3.0,5.2) | 2.6 (1.9,3.6) | 2.9 (2.7,3.0) | 3.0 (2.9,3.2) |
| **Diseases of the circulatory system** |  |  |  |  |  |  |  |
| Heart diseases | 268 | 461 | 0.5 (0.4,0.7) | 0.4 (0.2,0.9) | 0.3 (0.1,0.8) | 0.5 (0.5,0.6) | 0.5 (0.5,0.6) |
| Circulatory diseases | 290 | 358 | 0.5 (0.4,0.7) | 0.4 (0.2,0.9) | 0.6 (0.3,1.1) | 0.4 (0.3,0.4) | 0.4 (0.4,0.5) |
| **Diseases of the respiratory system** | 31,816 | 45,898 | 64.0 (62.3,65.7) | 59.7 (55.7,64.0) | 55.4 (51.7,59.2) | 50.0 (49.3,50.7) | 52.7 (52.1, 53.3) |
| **Diseases of the digestive system** | 7,534 | 9,762 | 12.4 (11.7,13.2) | 12.8 (11.1,14.9) | 10.8 (9.3,12.6) | 10.9 (10.6,11.2) | 11.2 (10.9,11.5) |
| **Diseases of the skin and subcutaneous tissue** | 3,330 | 3,954 | 4.7 (4.2,5.1) | 5.7 (4.6,7.2) | 4.5 (3.5,5.7) | 4.5 (4.3,4.7) | 4.5 (4.4,4.7) |
| **Diseases of the musculoskeletal system and connective tissue** | 805 | 1,040 | 1.1 (0.9,1.3) | 1.3 (0.8,2.1) | 1.5 (1.0,2.3) | 1.2 (1.1,1.3) | 1.2 (1.1,1.3) |
| **Diseases of the genitourinary system** | 3,482 | 5,436 | 6.1 (5.6,6.7) | 6.0 (4.8,7.5) | 5.6 (4.6,7.0) | 6.3 (6.0,6.5) | 6.2 (6.0,6.5) |
| **Certain conditions originating in the perinatal period** | 19,576 | 22,541 | 27.2 (26.1,28.3) | 28.2 (25.5,31.2) | 24.5 (22.1,27.1) | 25.4 (24.9,25.9) | 25.8 (25.3,26.2) |
| **Congenital malformation, deformations and chromosomal abnormalities** | 7,247 | 11,366 | 13.8 (13.0,14.6) | 12.8 (11.1,14.9) | 12.1 (10.4,13.9) | 12.9 (12.6,13.3) | 13.0 (12.7,13.4) |
| **Injury, poisoning, and certain other consequences of external causes** | 8,608 | 9,990 | 11.5 (10.8,12.3) | 14.3 (12.4,16.5) | 10.2 (8.7,12.0) | 11.4(11.1,11.8) | 11.5(11.2,11.8) |
| **Total** | 95,715 | 197,735 | 251.5<br>(248.2,255.0) | 264.1<br>(255.5,272.9) | 215.0<br>(207.7,222.5) | 220.8<br>(219.3,222.2) | 226.8<br>(225.5,228.1) |

Across all disease groups, the highest admission rates were for diseases of the respiratory system (52.9 per 1,000 child-years, 99% CI: 52.3 to 53.6), while the lowest were for mental and behavioural disorders (0.4, 0.4 to 0.5) and diseases of the circulatory system (0.5, 0.5 to 0.6 for heart diseases and 0.4, 0.4 to 0.5 for circulatory diseases).

Children exposed in utero during the wild-type period had higher admissions rates for malignant neoplasms (based on 195 disease cases), diseases of the blood and blood-forming organs and certain immune disorders, and injury, poisoning, and certain other consequences of external causes, but lower admission rates for benign neoplasms; while children exposed in utero during the Alpha period had lower admission rates for diseases of the respiratory system.

## Sensitivity analysis

After excluding 1,648 emergency admissions (1.1% of all first emergency admissions) and 101 planned admissions (0.2% of all first planned admissions) that were COVID-19 related, the sensitivity analysis restricted to non-COVID-19-related admissions showed results consistent with the main analysis (Figure C1 and C2 in Supplementary Sensitivity Analysis). Sensitivity analysis excluding admissions and deaths within the first 7 days of life, with follow-up beginning on day 8, also produced similar findings (Figure C3). Findings from logistic regression models were consistent with those from the Cox proportional hazard models used in the main analysis (Figure C4). When restricting the comparator group to children whose mothers tested negative during the same variant period as those who tested positive, all hazard ratios were slightly attenuated, and children exposed in utero during the Alpha period no longer showed a lower hazard of emergency admissions than the unexposed children (Figure C5 and Table C1). The overall conclusion remained unchanged.

## Discussion

### Key findings

Based on our England cohort of a quarter of a million children with a pregnancy start date in the first six months of the COVID-19 pandemic, 5% of children had a record of exposure to in utero SARS-CoV-2. We found no evidence of increased risk of either first emergency or planned admission up to 40 months of age associated with in utero exposure to SARS-CoV-2 during the wild-type and Alpha periods. Compared with unexposed children, children exposed in utero during the Alpha period had a slightly lower overall emergency admission rate, while no difference was observed for those exposed during the wildtype period. Within the exposed groups, no differences were found in the hazard of first emergency or planned admissions according to trimester of maternal SARS-CoV-2 infection. Diagnosis-specific admission rates showed modest differences, with some elevated rates of malignant neoplasms and injury-related admissions among children in utero exposed during the wild-type period, and blood and immune disorders among children in utero exposed during either period.

### Strengths and limitations

A key strength of this study is the use of multiple, national linked administrative health datasets, thereby ensuring representativeness and minimising loss to follow-up. The follow-up period of 40 months is longer than similar studies published to date. Our examination of both emergency and planned admissions capture any potential severe health outcomes requiring hospital admission. By defining the cohort based on the timing of conception rather than birth, we minimised fixed cohort bias[46,60]. We combined mother-baby linkage derived from HES[31] with those recorded in birth notification data[61] to maximise cohort coverage, and integrated information from various databases to minimise missing data. Analyses adjusted for key maternal and sociodemographic confounders, and multiple sensitivity analyses supported the robustness of the findings.

Our study has some limitations. First, although we used two COVID-19 surveillance datasets and hospital admission data to ascertain maternal infection status, we were unable to capture asymptomatic infections that were not tested or test-positive cases from the community that were not reported; those infections were included in the “no reported result” group. Underreporting may have been a particular issue during the wild-type period, when testing capacity was limited (February to April 2020) and community testing was not widespread (before 2020)[62–64]. In particular, since we found that children whose mothers had a negative test reported were less likely to live in deprived areas, but more likely to have chronic conditions recorded, some differential misclassification of infection status reflecting different testing and healthcare seeking behaviours among pregnant women is likely.

Second, in utero exposure was classified by the dominant circulating variant at the time of testing or diagnosis, as laboratory confirmation of variants was not available. However, given the high transmissibility and rapid dominance of the Alpha variant in the UK, the risk of variant misclassification is likely to be minimal[65–67]. Our findings are limited to the wildtype and Alpha period and may not be generalisable to later variants or to pregnancies in vaccinated women, as we focused on pregnancies prior to vaccine rollout to avoid any impact of vaccination on child health oucomes[68,69]. Further studies, including linked vaccination data, are required to confirm whether out findings are applicable to Delta and Omicron SARS-CoV-2 variants.

Third, our outcome measures were limited to hospital admissions. We were unable to access primary or outpatient care use or developmental or health outcomes not requiring hospital admissions.

Lastly, as the cohort was followed from birth rather than conception, findings are conditional on live births and do not account for the competing risks of stillbirth and pregnancy loss, the risk of which may increase by SARS-CoV-2 infection during pregnancy. For example, a population-based study of 0.8 million pregnancies in England and Wales showing an association between maternal COVID-19 diagnosis and stillbirth within 14 days of infection[70]. Further studies could integrate abortion and stillbirth data to fully describe the effect of SARS-CoV-2 infection during pregnancy.

### Comparison with other studies

Several studies have investigated health outcomes in neonates and infancy following in-utero exposure to SARS-CoV-2, but few have examined long-term, all-cause outcomes separately for emergency and planned hospital admissions using population-wide data. A small cohort study of 339 children (96 exposed and 243 unexposed) born in England and Wales between March 2020 and February 2021 - spanning the wild-type and Alpha variant periods - found higher admission rates by 21 months of age among those with perinatal exposure to SARS-CoV-2 (38% of exposed vs. 21% of unexposed children had at least one admission)[71]. However, it remains unclear whether these differences were driven by in utero exposure, neonatal infection, or both.

Though findings were not consistent across all studies, the majority of studies worldwide reported a higher risk of neonatal intensive care unit admission in babies exposed to SARS-CoV-2 in utero compared to unexposed babies, primarily due to conditions such as respiratory distress and hyperbilirubinemia[1,16,17,72–74]. This increased risk is thought to be largely driven by preterm delivery, often resulting from clinical decisions related to maternal health complications or medically induced preterm birth, rather than a direct viral effect on the foetus[75–78]. Our study, with a longer follow-up of 40 months, found no evidence of an increased overall hazard of hospital admission, particularly for diseases of the respiratory system, among children born to mothers who tested positive for SARS-CoV-2 during pregnancy compared with those born to test-negative mothers. Note that any admission within one day of the birth admission was also excluded, as part of our linkage of HES episodes into admissions, which may account for part of the discrepancy with earlier studies. Differences in findings may also reflect variation in follow-up duration and control group selection. Some previous studies[17,79] compared test-positive pregnancies with a broader reference group that included both test-negative women and those without any recorded test results during pregnancy – a heterogeneous group likely comprising true negatives, under-ascertained positives, and pregnant women with diverse health status, exposure risks, and testing and care-seeking behaviours. In contrast, we restricted our control group to test-negative pregnancies who actively engaged with testing, thereby reducing potential selection bias related to differential healthcare access and care-seeking behaviour.

Clear socio-demographic differences between the no-recorded-result and test-negative groups in our data indicate that it is not appropriate to assume the former were necessarily uninfected. Results related to the no-recorded-result group in this study should therefore be interpreted with caution.

Existing research on the impact of in utero exposure to SARS-CoV-2 has primarily focused on developmental delays in gross and fine motor, problem-solving, communication, and social skills, assessed with various scales[18,19,24,25,27,80–85]. However, these studies mostly had relatively short follow-up periods and reported mixed findings. A systematic review of 17 studies published up to June 2023 reported mild delays in gross and fine motor skills up to six months of age and in social and language domains between six and 12 months[80]. In contrast, three subsequent studies - including two prospective cohorts from Canada and the US with a 24-month follow-up period[24,26] and a Scottish population-based retrospective cohort with a follow-up period of 13 to 15 months[25] – found no association between in utero exposure to SARS-CoV-2 and developmental concerns. Unlike these studies, our analysis was limited to hospital admissions and therefore only captured severe cases of neurological disorders requiring hospital admission, making direct comparisons difficult.

This study observed a lower hazard of emergency hospital admission among children with in utero exposure during the Alpha period compared to those with no exposure. One reason for observing this difference during the Alpha, but not wild-type, period may be that our comparator group (test-negative) likely included symptomatic women tested in the community, particularly during the wild-type period when testing was limited to symptomatic individuals. This may represent a less healthy comparator group than that included during the Alpha period when broader community testing was in place. Further, during the wild-type period, limited testing likely captured more severe infections or high-risk pregnancies, while broader testing during the Alpha period identified a more heterogeneous group of possibly healthier exposed pregnancies. In the sensitivity analysis using children whose mothers tested negative during the same variant period as those who tested positive as the comparator group, the lower hazard of emergency admissions among children in utero exposed during the Alpha period no longer exists. Whether neonatal outcomes differ by variant following in utero exposure remains inconsistent and evidence comparing different variants is limited[50,61,86–89]. For example, research from Italy and Scotland showed a higher risk of premature births[90,91], small-for-gestational age[91], and low Apgar score[90] and evidence from France, Switzerland[61], and Malawi[92] showed higher stillbirth rates associated with the Delta variant, while a US study showed no difference in Apgar scores, NICU admissions or need for respiratory support between the pre-Delta and Delta variants, although with longer hospital stay in neonates born during the Delta period[87].

Our study, focusing on longer-term outcomes after birth, found broadly similar crude admission rates between exposed and unexposed children across most disease groups.

However, children exposed in utero during the wild-type period had higher crude admission rates for malignant neoplasms and diseases of the blood and blood-forming organs, and certain disorders involving the immune mechanism. We did not observe this finding for children exposed to SARS-CoV-2 in utero during the Alpha wave. We consider it unlikely to be due to SARS-CoV-2 exposure during pregnancy, but instead may reflect changes in healthcare delivery for children with cancer earlier in the pandemic, or potentially in exposure to infections in early life (which has been associated with leukaemia risk[93–96]) due to the changes in seasonality of common infections seen during and after the COVID pandemic. These impacts may well be differential by exposure group, due to the complex changes in SARS-CoV-2 circulation, testing availability, and healthcare availability over time. The concurrent increase in admission rates for injury, poisoning, and certain other consequences of external causes, along with the absence of malignant neoplasm cases among children exposed in utero during the Alpha period, raises the possibility of unadjusted confounding between exposure groups. Further studies, including linkage to cancer registration data to obtain the date of cancer diagnosis, rather than the date of hospital admission with cancer diagnosis recorded, are required.

Our study shows no difference in the hazards of first admission in early childhood by trimester of in-utero exposure, after adjusting for the variants exposed and other maternal and sociodemographic confounders. However, interpretation of the potential impact of the first-trimester exposure should be cautious, as the number of cases exposed in early pregnancy was small and confined to the wild-type period. Pregnancies affected in the first trimester may be more likely to result in early pregnancy loss, which is not captured in our live-birth cohort and may lead to underestimation of adverse outcomes associated with early pregnancy exposure. Despite these limitations, our findings align with most previous studies that reported no consistent evidence of trimester-specific effects on a range of perinatal and early childhood outcomes[26,71,97].

### Clinical and policy implications

Overall, findings from this study suggest no evidence for increased risk of emergency or planned hospital admission in children with in utero SARS-CoV-2 exposure during the wild-type and Alpha period up to 40 months of age, providing reassurance to families affected by maternal COVID-19 infection in the early pandemic period. These findings support ongoing public health messaging that maternal SARS-CoV-2 infection, at least in its earlier variants, does not indicate an increased risk of children exposed in utero requiring hospital admission. Further studies with longer follow-up are needed to assess the health consequences of in utero SARS-CoV-2 exposure with onset in later childhood or adulthood, ideally accounting for survivorship bias.

## Conclusion

In conclusion, our study, the largest population-based cohort with the longest follow-up period, found no evidence for an increase in children’s emergency or planned admission rates up to 40 months of age, following in utero SARS-CoV-2 exposure during the wild-type and Alpha periods. No differences in either emergency or planned admissions rates were observed across trimesters of exposure. Further studies should examine disease-specific outcomes and in utero exposure to SARS-CoV-2 during later variant periods.

## Data Supplement

Supplementary Methods

Supplementary Tables and Figures

Supplementary Sensitivity Analysis

Supplementary STROBE Checklist

## Data Availability

This study used NHS Hospital Episode Statistics data which was provided within the terms of a data sharing agreement (DARS-NIC-393510-D6H1D-v9.3) by NHS England.

## Acknowledgement

We acknowledge funding from the European Union’s Horizon Grant and support from the SARS-CoV-2 variants Evaluation in Pregnancy and Paediatrics cohorts (VERDI) project. This work used data provided by the public and patients and collected by the NHS as part of their care and support. NHS Hospital Episode Statistics data was provided within the terms of a data-sharing agreement (DARS-NIC-393510-D6H1D-v9.3) to the researchers by NHS England. We acknowledge the help and support of Dr. Linda Wijlaars and Professor Darren Hargrave.

## Contributor and guarantor information

PH, CT, LW, and ML contributed to the development and conduct of the study. ML, LW, and FA contributed to the data cleaning and analyses. ML wrote the first draft of the manuscript with contributions from PH and CT. All authors edited drafts of the manuscript and approved the final version of the article. The corresponding author attested that all listed authors meet authorship criteria and that no others meeting the criteria have been omitted. Pia Hardelid is the guarantor.

## Data sharing

The data do not belong to the authors and may not be shared by the authors, except in aggregate form for publication. The data is provided by patients and collected by the NHS as part of their care and support. Data can be obtained by submitting a data request through the NHS England Data Access Request Service.

## Transparency statement

The lead author affirms that the manuscript is an honest, accurate, and transparent account of the study being reported; that no important aspects of the study have been omitted; and that any discrepancies from the study as planned (and, if relevant, registered) have been explained.

## Dissemination to participants and related patient and public communities

Dissemination to participants is not possible as this study analysed pseudonymised data only. Dissemination to women, families, and healthcare practitioners will be undertaken via social media and the programme website (https://verdiproject.org/) and through summary articles for professional and third-sector organisations.

This is an Open Access article distributed in accordance with the terms of the Creative Commons Attribution (CC BY 4.0) license, which permits others to distribute, remix, adapt and build upon this work, for commercial use, provided the original work is properly cited. See: http://creativecommons.org/licenses/by/4.0/.

