## supplementary appendix for "Impact of in utero exposure to SARS-CoV-2 on children’s hospital admission: National birth cohort study of 262,000 children in England"

### Supplementary Methods

#### Data sources and identification of birth cohort

Birth notifications are reported by the attending midwife or other birth attendants within 36 hours of birth to the NHS[1]. They contain information about the newborn (e.g., birth weight and ethnicity) and mother's HES unique identifier (Token ID), through which mothers were linked to their babies in the birth cohort. HES APC is collated by NHS England and covers all hospital admissions paid for by the English NHS[2]. The HES mother-baby linked dataset, developed by Feng et al., based on birth and delivery records identified from HES APC[2,3], was used to identify mothers for births recorded solely in HES.

Birth admissions were determined using the criteria developed by Zylbersztejn et al.[4], based on diagnostic and procedure codes, healthcare resource group codes and administrative variables from hospital admissions within one month of life (Table A1). If more than one birth admission was identified for a child, the first one was seen as the sole birth record. Birth admissions were excluded from the outcome ascertainment.

**Table A1. Criteria for identifying birth admission in HES**

| | Variable used | Inclusion Criteria<br>(value recorded in HES and explanation) |
|---|---|---|
| **Diagnostic codes (ICD-10)** | | Z38: Liveborn infants according to place of birth and type of delivery<br>Z37: Outcome of delivery |
| **Healthcare Resource Group Codes** | **Version 3.5** | N01: Neonates - Died <2 days old<br>N02: Neonates with Multiple Minor Diagnoses<br>N03: Neonates with one Minor Diagnosis<br>N04: Neonates with Multiple Major Diagnoses<br>N05: Neonates with one Major Diagnosis |
| | **Version 4.0 (in use since financial year 2011/12)** | PB01Z: Major Neonatal Diagnoses<br>PB02Z: Minor Neonatal Diagnoses<br>PB03Z: Healthy Baby |
| **HES Specific Fields** | **Episode type** *(epitype)* | 3: Birth episode<br>6: Other birth event |
| | **Patient classification** *(classpat)* | 5: Mothers and babies using only delivery facilities |
| | **Admission method** *(admimeth)* | 82: Other: babies born in health care provider<br>83: Other: babies born outside the health care provider, except when born at home as intended<br>2C: Baby born at home as intended (available from 2013/14) |
| | **Neonatal Care** *(neocare)* | 0: Normal care<br>1: Special care<br>2: Level 2 intensive care (high dependency intensive care<br>3: Level 1 intensive care (maximal intensive care) |

*HES, Hospital Episode Statistics; ICD-10, International Statistical Classification of Diseases and Related Health Problems. Financial years in England run from 1st April to 31st March the following year. Healthcare resource groups, developed by National Casemix Office, are standard grouping of clinically similar treatments which use common levels of healthcare resource.*

Due to confidentiality restrictions, birth notifications contain the month of birth rather than the exact date. The date of birth was estimated by the birth admission date, the earliest admission date, or the 15th day of the recorded birth month. If a birth admission was identified in the HES APC using criteria listed in Table A1, the birth admission date was used as the estimated date of birth. If a birth admission was not identified but there is an admission in the same month of birth, the admission date was used as the estimated date of birth. If a birth admission was not identified in HES APC and there was no recorded admission in the same month of birth, the 15th day of the recorded birth month was used as the estimated date of birth.

In the study birth cohort, 97.4% of births appear in both HES and birth notifications, 2.4% appear in birth notification only, and 0.2% appear in HES only (Figure A1).

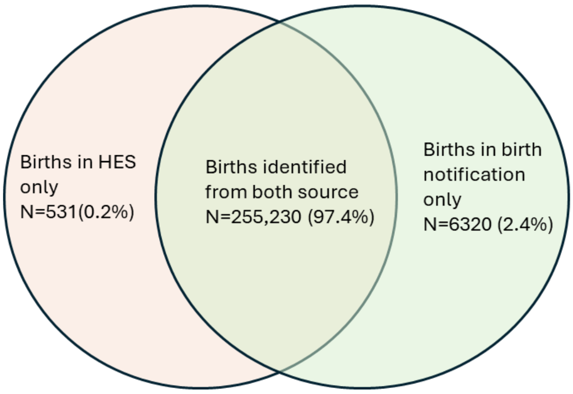


**Figure A1. Source of births identified from Hospital Episode Statistics and birth notifications**

Mothers were linked to their babies using the linkage in the birth notifications, and then the linkage developed by Feng et al. if the birth was identified in HES only[2,3]. Birth notifications contain a variable indicating the mother's TokenID (unique identifier). In the study cohort, the agreement between Qi's mother-baby link and birth notifications was 99.2%.

Death registrations are collected by the Office for National Statistics (ONS) as part of the national vital registration systems[5].

#### Cleaning and linking hospital admissions

We extracted all episodes of care for babies identified in the study birth cohort. We removed episodes with no clinical information recorded (e.g., unfinished episodes). We then cleaned and validated date variables (such as admission and discharge dates) which can contain recording errors (e.g., if the recorded admission date was after the discharge date). Finally, we de-duplicated the episodes. Details of data cleaning rules are described in Table A2.

We then linked episodes into admissions using an algorithm developed by Hardelid et al.[6] An admission was defined as a continuous period of time that a child spent under NHS hospital care. Hospital transfers and admissions within 1 day of each other were treated as one inpatient admission. Lastly, we derived the most commonly recorded value of region of residence, IMD score and post code per admissions. Stata codes are published on [UCL Child Health Informatics Group GitHub site].

**Table A2. Cleaning rules for HES longitudinal records**

| Criterion | | Action |
|---|---|---|
| **Drop episodes with no clinical information recorded** | Unfinished episodes | Drop (as usually more complete record was available) |
| | Missing episode end date | Drop (as usually more complete record was available) |
| | Only clinical information recorded was diagnosis "R69" – "*Illness, unspecified*" | Drop (as usually more complete record was available) |
| | No recorded diagnoses | Drop (as usually more complete record was available) |
| **Validate and correct date variables (admission and discharge dates, episode start and end dates)** | Admission date missing | Replace to episode start date for the first episode of the admission (*epiorder*=1)<br>Else, replace to admission date from episode with episode order smaller by 1 and closest episode start date<br>Else, replace with episode start date |
| | Episode start date missing | Replace to admission date for the first episode of the admission (*epiorder*=1)<br>Else, replace to episode end date from another episode with the same admission date and lower episode order |
| | Episode end date missing | Removed as part of exclusion criteria |
| | Episode start> episode end | Replace episode start with admission date if the issue is with the recording of episode start (episode start > episode end ≥ admission date)<br>Replace episode end with episode start date if the issue is with the recording of episode end (episode start = admission date > episode end)<br>Switch episode start with episode end, and admission date with discharge date if they were incorrectly recorded (episode start > episode end & admission date > discharge date where discharge date is not missing) |
| | Admission date > episode start | Replace episode start date with admission date |
| | Admission date > episode end | Replace episode end date with episode start date |
| **Validate and correct date variables (admission and discharge dates, episode start and end dates)** | Discharge date missing | Discharge date is recorded only on the last episode of care. Therefore, I generated a maximum discharge date by HESID and admission date as the "complete" discharge date.<br>If "complete" discharge date was missing (when discharge date was not recorded for an admission), I replaced it with maximum episode end date by HESID and admission date.<br>If "complete" discharge date was smaller than the maximum episode end date, I replaced it with the maximum episode end date. |

| | | |
|---|---|---|
| | Episode ends in a different year than it starts | Drop if the difference is greater than or equal to two. It seems impossible to be seen by only one consultant while staying in the hospital for 2 years so it must be a data error. |
| | Missing episode start age | No such observations |
| | Missing episode end date | Generate an age using episode start and end dates for episodes with startage=7001 (“less than 1 day”) |
| | Age at start of episode > age at end of episode | Switch age at start with age at end of episode |
| | Epistart – Epiend > 365 | Episodes that lasted more than 1 year were assumed to be recording errors and dropped as it is unlikely that a patient would be seen by just one consultant for that long. |
| **Drop duplicates** | Exact duplicates | Drop duplicates in terms of: HESID, age at start and end of admission, month and year of birth, gender, post code, start and end date of the episode, episode order, admission and discharge dates, provider code,all diagnoses and operations and cause of injury |

*HES, Hospital Episode Statistics; IMD, Index of Multiple Deprivation.*

#### Identifying stillbirths and multiple births

**Table A3. Rules for identifying stillbirths and multiple births in Hospital Episodes Statistics**

| Stillbirths | Rules |
|---|---|
| Multiple births | ICD-10 codes (diag_XX variables) include any of Z383, Z384, Z385, Z372, Z373, Z375, Z376, Z377, Z386, Z387 |
| | Birth order (‘birordr’ variable)>1 |
| | Number of babies (‘numbaby’ variable)>1 |
| Still births | ICD-10 codes (diag_XX variables) include any of Z371, Z373, Z374, Z376, Z377, P95 |
| | Discharge method (‘dismeth’ variable) is 5-stillbirth |
| | Birth status (‘birstat’ variable) is 2-still birth ante-partum, 3-still birth intra-partum, or 4-still birth indeterminate |

#### Identifying planned and emergency admissions

The type of admissions was determined by the “admimeth” variable (method of admission) in HES APC, based on the classification in Table A4. The admission method of the first episode of each admission was considered the admission method of the admission. Transfer admission, birth admission, maternity admissions, and admissions with unclear admissions were excluded from the analysis.

**Table A4. Definition, frequency, and inclusion of each type of admissions**

| Type of admission | Value in “Admimeth” variable in HES APC | Inclusion |
|---|---|---|
| Planned admission | 11, 12, 13 | Included in analysis of planned admission |
| Emergency admission | 21, 22, 23, 24, 25, 28, 2A, 2D | Included in analysis of emergency admission |
| Transfer admission | 2B,81 | Excluded |
| Birth admission | 2C, 82, 83 | Excluded |
| Maternity admission | 31, 32 | Excluded |
| Unclear | 98, 99 | Excluded |

#### Cleaning multiple SARS-CoV-2 tests on the same day

Pillar 1 data contains only positive PCR tests. Pillar 2 contains both PCR and rapid lateral flow tests with positive, negative, or unknown results. In Pillar 2, the variable "testresult" was used to determine the type of test and the results (Table A5). If there was more than one test result on the same day, rules in Table A6 were used to reduce them to one final result per day. Results from PCR tests were prioritised when test results were available in both PCR and lateral flow testing on the same day, as PCR has higher sensitivity than antigen tests[7,8].

**Table A5. Determining the test type and results in Pillar 2 dataset**

| Value in "testresult" variable | Type of test | Test result |
|---|---|---|
| SCT: 1240581000000104 | PCR | Positive |
| SCT: 1240591000000102 | PCR | Negative |
| SCT: 1321691000000102 | PCR | Unknown |
| SCT: 1322781000000102 | LFT | Positive |
| SCT: 1322791000000100 | LFT | Negative |
| SCT: 1322821000000105 | LFT | Unknown |

**Table A6. Rules of reducing multiple SARS-CoV-2 tests on the same day to one final result per day**

| Scenarios | SARS-CoV-2 test results for the day |
|---|---|
| 1) One test or multiple tests with consistent result | Use the consistent results |
| 2) Multiple tests with inconsistent results but PCR test results are consistent | Use the consistent PCR results |
| 3) Multiple LFT tests with inconsistent results and no PCR test result | If any of the LFT test result is positive, treat as positive; if LFT test results are either negative or unknown, treat as negative |
| 4) Multiple PCR tests with inconsistent results | If any of the PCR tests is positive, treat as positive; if PCR test results are either negative or unknown, treat as negative |

#### Calculating rate of emergency admission and planned admissions

All emergency and planned admissions (not just the first) were included in the calculation of admission rates, which were estimated separately for emergency and planned admissions. When calculating emergency admission rates, person-time at risk excluded periods of hospital stay during emergency admissions but included time spent in hospital for planned admissions; vice versa for planned admission rates. To account for multiple testing, 99% confidence intervals were reported alongside admission rates.

#### Directed Acyclic Graph

The exposure of this study is the recorded exposure to SARS-CoV-2 in utero, and the outcome of interest is child hospital admissions. Based on the directed acyclic graph[9] (Figure A1), the minimal sufficient adjustment sets contain area deprivation,  ethnicity, maternal age, maternal chronic conditions, and the year-month of conception.

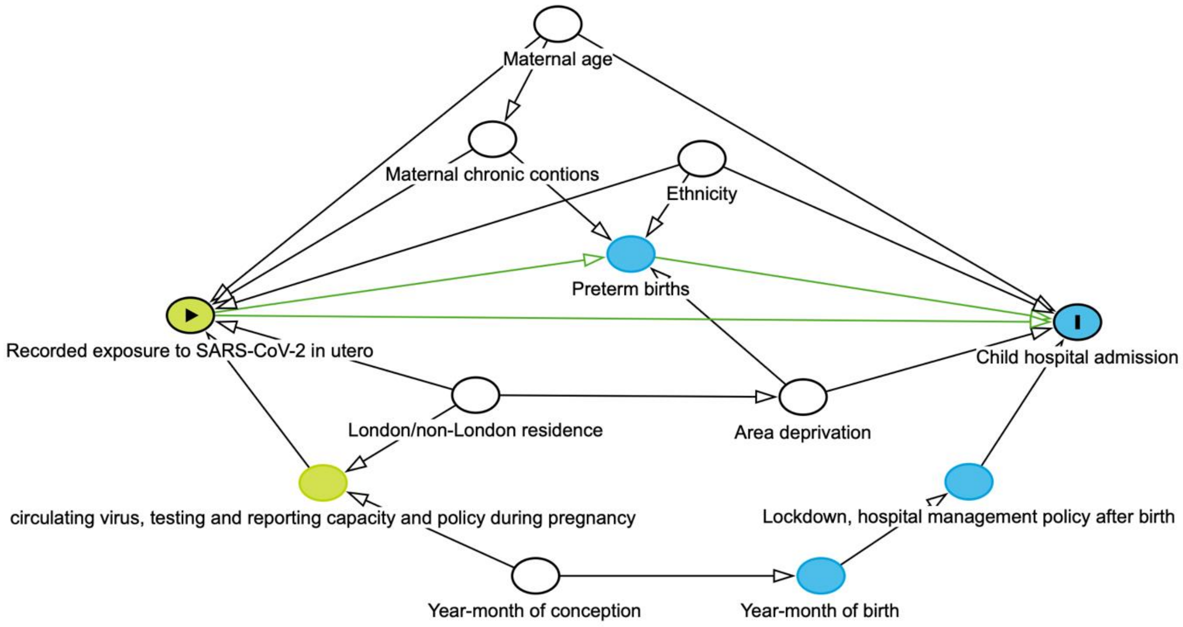


**Figure A2. Directed Acyclic Graphs to identify cofounders**
Note: Residence in London at birth was specifically adjusted for, as the transmission of SARS-CoV-2 was different in London[10–12], and hospital admission rates for young children are lower in London compared to the rest of England[13,14].

#### Categorisation of primary diagnosis

**Table A7. Categorising primary diagnosis into disease groups**

| Disease group | ICD-10 codes in the primary diagnosis of the first episode of the admission |
|---|---|
| **Certain infectious and parasitic diseases** | |
| Bacterial infectious diseases | A00-A99 |
| Non-bacterial infectious diseases | B00-B99 |
| **Neoplasms** | |
| Malignant neoplasms | C00-C97 |
| Benign neoplasms | D00-D49 |
| **Diseases of the blood and blood-forming organs and certain disorders involving the immune mechanism** | D50-D89 |
| **Endocrine, nutritional and metabolic diseases** | E00-E90 |
| **Mental and behavioural disorders** | F00-F99 |
| **Diseases of the nervous system** | G00-G99 |
| **Diseases of the eye and adnexa** | H00-H59 |
| **Diseases of the ear and mastoid process** | H60-H95 |
| **Diseases of the circulatory system** | |
| Heart diseases | I00-I52 |
| Circulatory disease | I60-I99 |
| **Diseases of the respiratory system** | J00-J99 |
| **Diseases of the digestive system** | K00-K93 |
| **Diseases of the skin and subcutaneous tissue** | L00-L99 |
| **Diseases of the musculoskeletal system and connective tissue** | M00-M99 |
| **Diseases of the genitourinary system** | N00-N99 |
| **Certain conditions originating in the perinatal period** | P00-P96 |
| **Congenital malformations, deformations and chromosomal abnormalities** | Q00-Q99 |
| **Injury, poisoning and certain other consequences of external causes** | S00-S99, T00-T98 |

Note: Disease categorisation was developed by Hviid et al.[21] based on the International Statistical Classification of Diseases and Related Health Problems, $10^{th}$ Revision

### Supplementary Tables and Figures

#### Testing policy and criteria for Pillar 1 and Pillar 2

**Table B1. Who are included in each exposure group over study period**

| Exposure groups | Apr-Nov 2020 | Dec 2020-Mar 2021 | Apr-May 2021 |
|---|---|---|---|
| **Test-negative (all from community setting)** | Symptomatic individuals | Symptomatic individuals, asymptomatic key workers or individuals from high-risk areas | Everyone, regardless of symptoms |
| **Positive: Wild-type (Feb- mid-Dec 2020)** | Hospital: Symptomatic or asymptomatic cases<br>Community: Symptomatic cases | - | - |
| **Positive: Alpha (mid-Dec 2020-May 2021)** | - | Hospital: Symptomatic or asymptomatic cases<br>Community: Symptomatic individuals, asymptomatic cases of key workers or individuals from high-risk areas | Hospital: Symptomatic or asymptomatic cases<br>Community: Symptomatic or asymptomatic cases from any group of areas |
| **No-recorded-result** | Uninfected (not tested, or tested negative and not reported), or positive and not reported, or infected and not tested | | |

#### Number of SARS-CoV-2 tests and COVID-19 diagnoses over study period

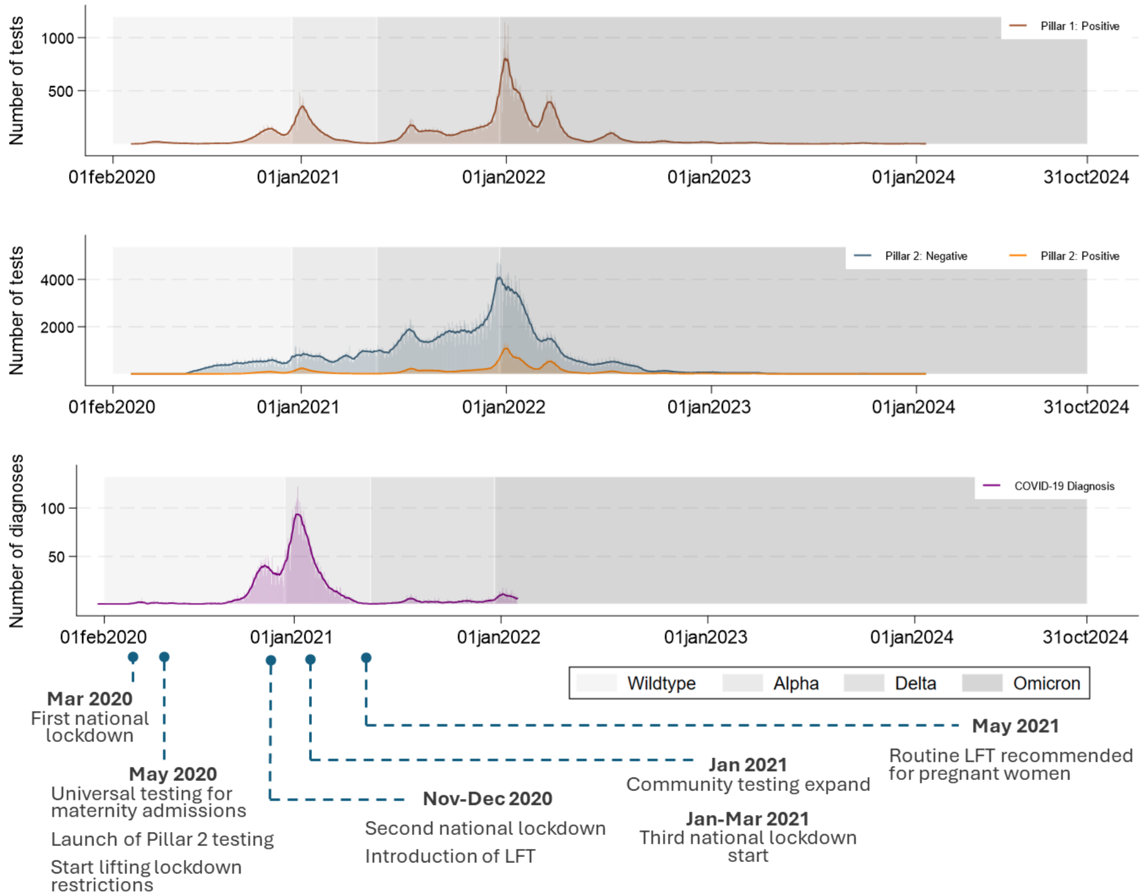


**Figure B1. Distribution of SARS-CoV-2 tests and COVID-19 diagnosis among mothers of the study cohort. From top panel to bottom panel: positive tests from Pillar 1; positive and negative tests from Pillar 2; positive diagnosis in HES.** Transparent bars show daily counts, and solid lines show the 15-day rolling average of daily counts (a week before + the index day + a week after). All recorded tests from mothers of the birth cohorts were included (including those taken postnatally) to illustrate changes in testing uptake over calendar time.

#### Kaplan-Meier curves

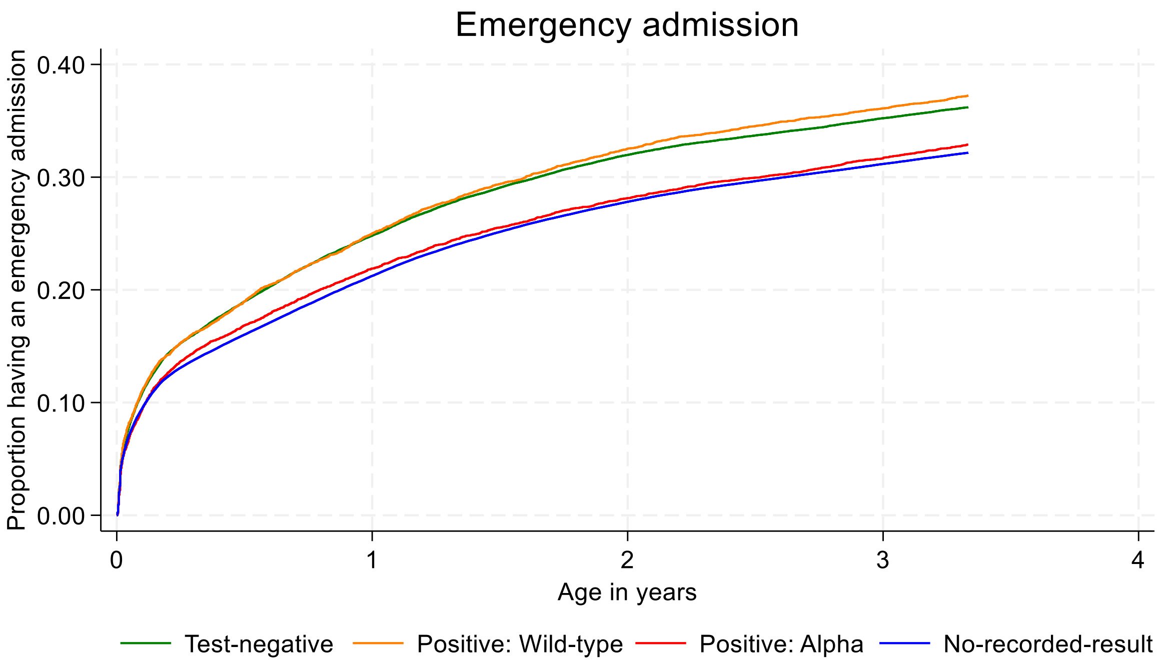


**Figure B2. Kaplan-Meier plot for children's first emergency admissions up to 40 months old by SARS-CoV-2 exposure in utero**

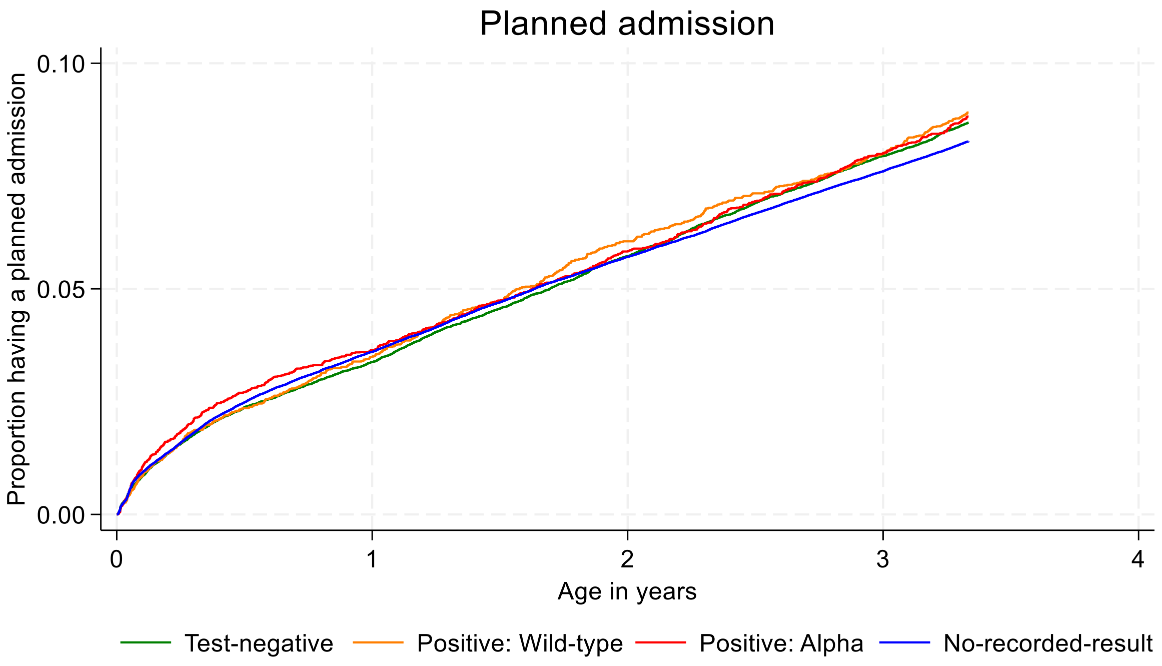


**Figure B3. Kaplan-Meier plot for children's first planned admission up to 40 months years old by SARS-CoV-2 exposure in utero**

**Table B2. Time to child's first emergency and planned admission up to 40 months old: crude and adjusted hazard ratio (99% confidence interval) by in utero exposure to SARS-CoV-2 during pregnancy, based on 257,241 children conceived between February and July 2020 in England**

| Exposure and covariates | Emergency admissions (99% CI) | | Planned admissions (99% CI) | |
|---|---|---|---|---|
| | Crude | Adjusted | Crude | Adjusted |
| **In-utero SARS-CoV-2 exposure** | | | | |
| Test-negative | 1 | 1 | 1 | 1 |
| Positive: Wild-type | 1.04 (0.98,1.09) | 1.03 (0.97,1.09) | 1.04 (0.93,1.16) | 1.05 (0.93,1.17) |
| Positive: Alpha | 0.89 (0.84,0.94) | 0.92 (0.87,0.97) | 1.01 (0.91,1.13) | 1.00 (0.90,1.12) |
| No-recorded-result | 0.86 (0.84,0.88) | 0.88 (0.86,0.90) | 0.95 (0.91,1.00) | 0.95 (0.90,0.99) |
| **Ethnicity** | | | | |
| White | | 1 (1.00,1.00) | | 1 (1.00,1.00) |
| Asian | | 0.98 (0.95,1.00) | | 0.89 (0.84,0.94) |
| Black | | 0.84 (0.80,0.88) | | 1.06 (0.97,1.15) |
| Mixed | | 0.91 (0.86,0.97) | | 0.93 (0.83,1.04) |
| Other | | 0.90 (0.86,0.95) | | 0.97 (0.89,1.06) |
| **Area deprivation** | | | | |
| Most deprived 10% | | 1 | | 1 |
| More deprived 10-20% | | 0.99 (0.95,1.02) | | 0.99 (0.92,1.06) |
| More deprived 20-30% | | 0.95 (0.91,0.98) | | 0.97 (0.91,1.04) |
| More deprived 30-40% | | 0.94 (0.91,0.98) | | 0.98 (0.91,1.05) |
| More deprived 40-50% | | 0.96 (0.92,0.99) | | 0.96 (0.89,1.04) |
| Less deprived 40-50% | | 0.92 (0.88,0.95) | | 0.92 (0.85,1.00) |
| Less deprived 30-40% | | 0.92 (0.89,0.96) | | 0.95 (0.88,1.03) |
| Less deprived 20-30% | | 0.93 (0.90,0.97) | | 0.95 (0.88,1.03) |
| Less deprived 10-20% | | 0.93 (0.90,0.97) | | 0.92 (0.88,0.99) |
| Least deprived 10% | | 0.86 (0.83,0.90) | | 0.95 (0.87,1.03) |
| **Mother's history of chronic disease** | | | | |
| No | | 1 | | 1 |
| Yes | | 1.33 (1.28,1.39) | | 1.34 (1.24,1.46) |
| **Maternal age (years)** | | | | |
| <25 | | 1 | | 1 |
| 25-29 | | 0.91 (0.89,0.94) | | 1.00 (0.94,1.06) |
| 30-34 | | 0.84 (0.82,0.86) | | 0.98 (0.93,1.04) |
| 35-39 | | 0.80 (0.77,0.82) | | 1.00 (0.94,1.07) |
| ≥40 | | 0.81 (0.77,0.85) | | 0.98 (0.89,1.08) |
| **London residence** | | | | |
| No | | 1 | | 1 |
| Yes | | 0.71 (0.69,0.73) | | 1.19 (1.13,1.25) |
| **Month of conception** | | | | |
| Feb 2020 | | 1 | | 1 |
| Mar 2020 | | 1.03 (1.00,1.06) | | 1.01 (0.95,1.08) |
| Apr 2020 | | 1.04 (1.01,1.08) | | 0.96 (0.90,1.02) |
| May 2020 | | 1.03 (1.00,1.06) | | 0.94 (0.89,1.00) |
| Jun 2020 | | 1.03 (1.00,1.06) | | 0.98 (0.92,1.04) |
| Jul 2020 | | 1.05 (1.02,1.08) | | 1.00 (0.94,1.07) |

#### Check for the proportional hazard assumption

The proportional hazard assumption has been assessed by including an interaction term between the exposure status to SARS-CoV-2 in utero and the time variable in the Cox proportional hazard model (age of the child), with adjustment for maternal ethnicity, area deprivation decile, mother's history of chronic conditions, maternal age, London residence, and month of conception. Likelihood ratio tests were used to compare the models with and without the interaction.

**Table B3. P-value from tests for the proportional hazard assumption**

| Interaction with age | Emergency admissions | Planned admissions |
|---|---|---|
| Positive: Wild-type | 0.401 | 0.974 |
| Positive: Alpha | 0.261 | 0.178 |
| No-recorded-result | 0.533 | <0.001 |
| Likelihood ratio test comparing with the model without the interaction between exposure status and time variable | 0.627 | <0.001 |

### Supplementary Sensitivity Analysis

#### Excluding SARS-CoV-2-related admissions

We identified SARS-CoV-2-related admissions using criteria defined by Hardelid et al in a previous study[22]: (i) an individual had tested positive for SARS-CoV-2 up to 14 days prior to hospital admission, on the day of admission, or in between the hospital admission and discharge date, and/or (ii) an International Classification of Disease-version 10 (ICD-10) diagnostic code for COVID-19 (U07.1-U07.2) had been recorded during admission as primary or secondary diagnosis. We cleaned and identified children's SARS-CoV-2 positive tests in the same way as the mother's SARS-CoV-2 tests, described in the method section of the main text.

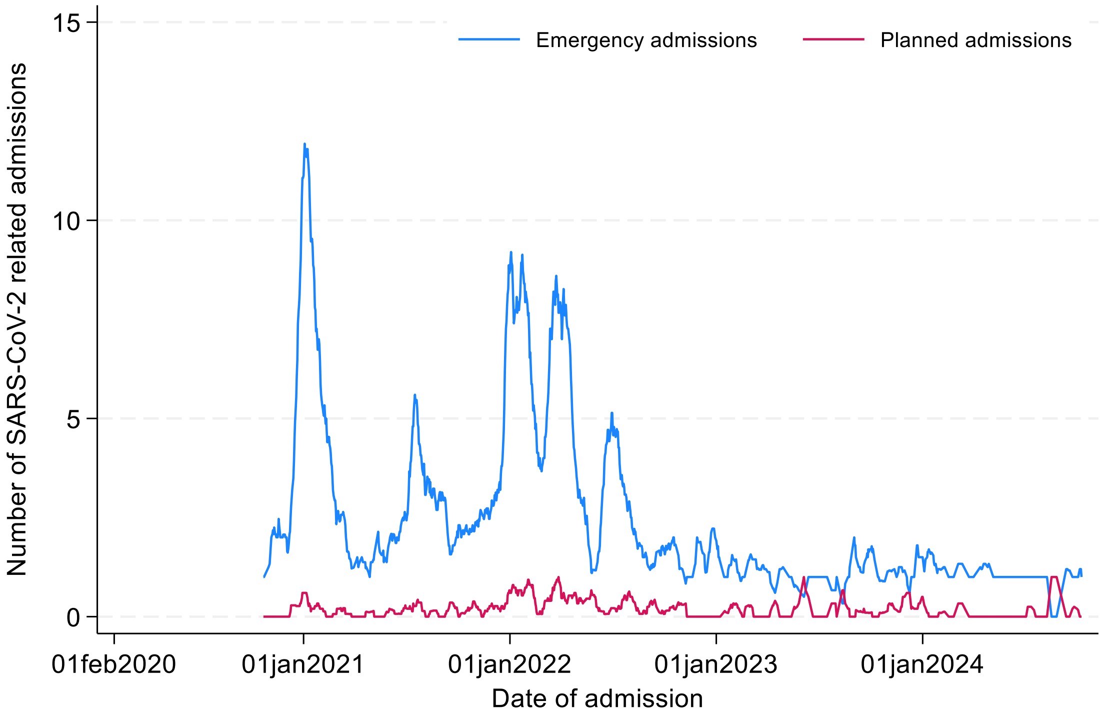


**Figure C1. The 15-day rolling average count (a week before + the index day + a week after) of SARS-CoV-2-related emergency admissions and planned admissions in the study cohort**

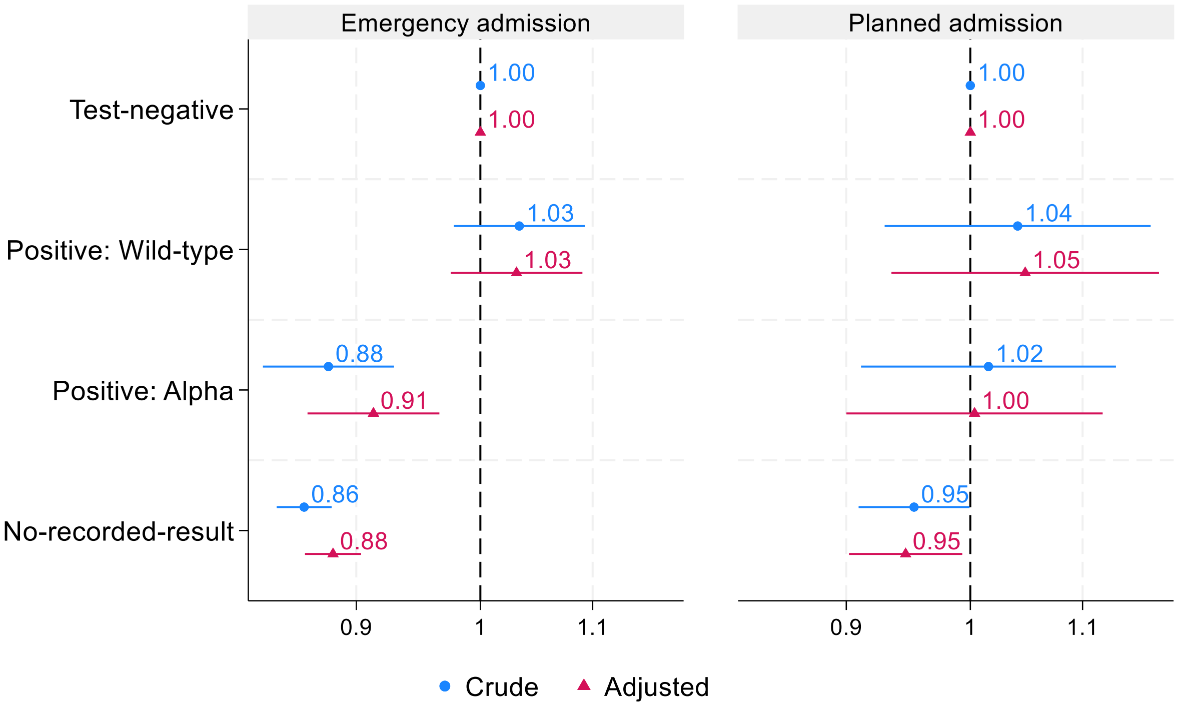


**Figure C2. Time to child's first non-COVID-19-related emergency and planned admission up to 40 months old: hazard ratio and 99% confidence interval by in utero exposure to SARS-CoV-2 during pregnancy, children conceived between February and July 2020 in England. Reference=Test-negative group**

Note: Results based on Cox proportional hazard regression models adjusted for maternal ethnicity, area deprivation decile, maternal chronic conditions, maternal age, London residence, and month of conception. A total of 257,241 children with complete data for all covariates were used.

#### Excluding admissions and deaths within the first 7 days of life, and starting follow-up from day 8

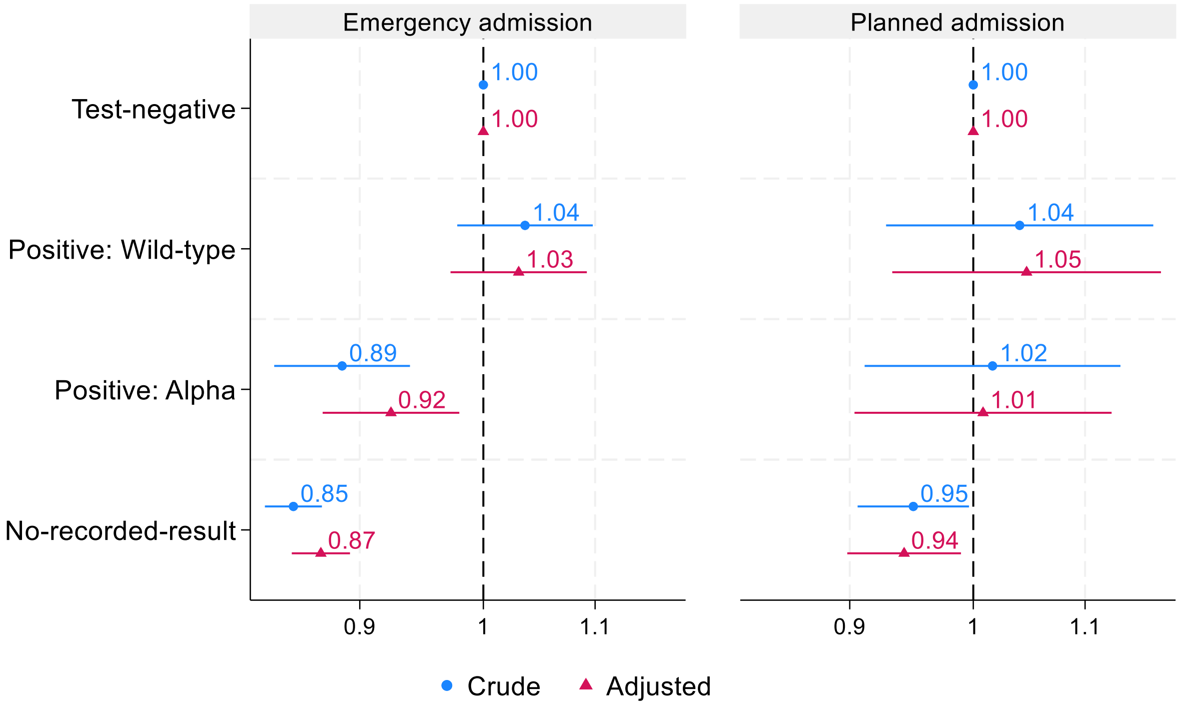


**Figure C3. Time to child's first emergency and planned admission between day 8 of life and 40 months old: hazard ratio and 99% confidence interval by in utero SARS-CoV-2 exposure status, children conceived and born between February 2020 and April 2021 in England. Reference=Test-negative group**

Note: Admissions and deaths within the first 7 days of life were excluded. Follow-up starts from day 8. Models adjusted for maternal ethnicity, area deprivation decile, maternal chronic conditions, maternal age, London residence, and month of conception. A total of 257,241 children with complete data for all covariates were used.

#### Logistic regression

Logistic regression shows similar results to those from Cox proportion hazard models.

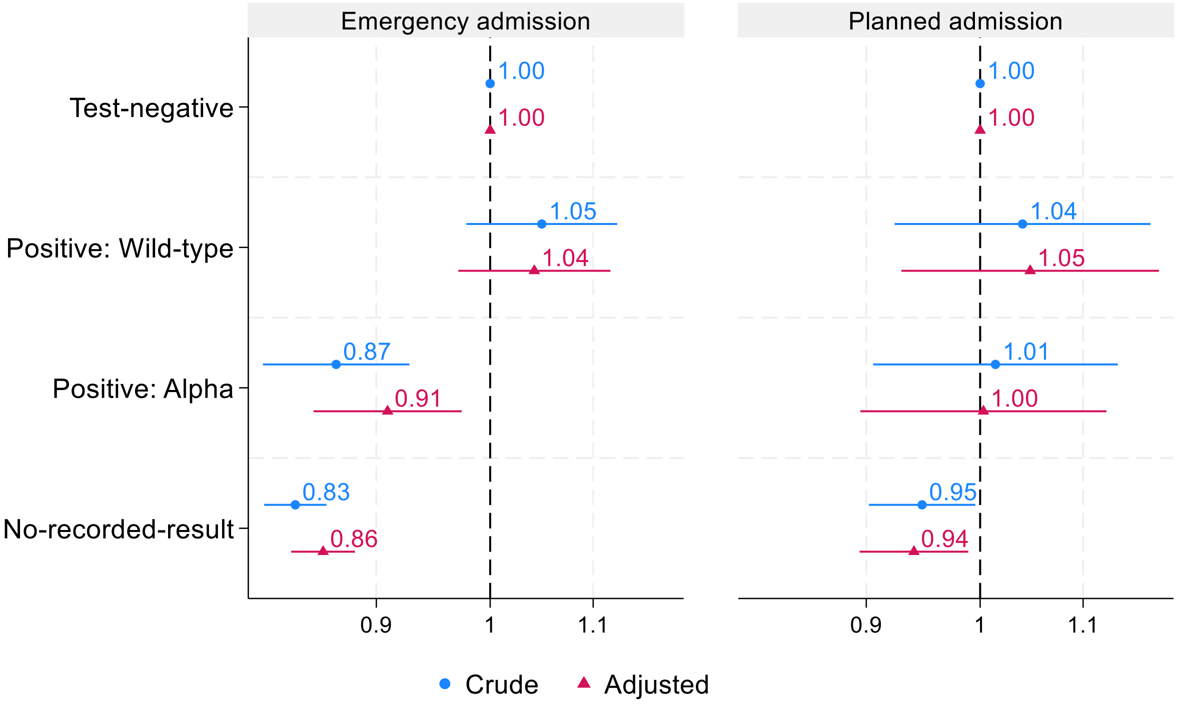


**Figure C4. Odds ratio (99% CI) for children's hospital admissions up to 40 months old according to SARS-CoV-2 exposure in utero, children conceived between February and July 2020 in England. Reference=Test-negative group**

Note: Results based on Logistic regression models adjusted for maternal ethnicity, area deprivation decile, maternal chronic conditions, maternal age, London residence, and month of conception. A total of 257,241 children with complete data for all covariates were used.

#### Separate control (test-negative group) by variant period

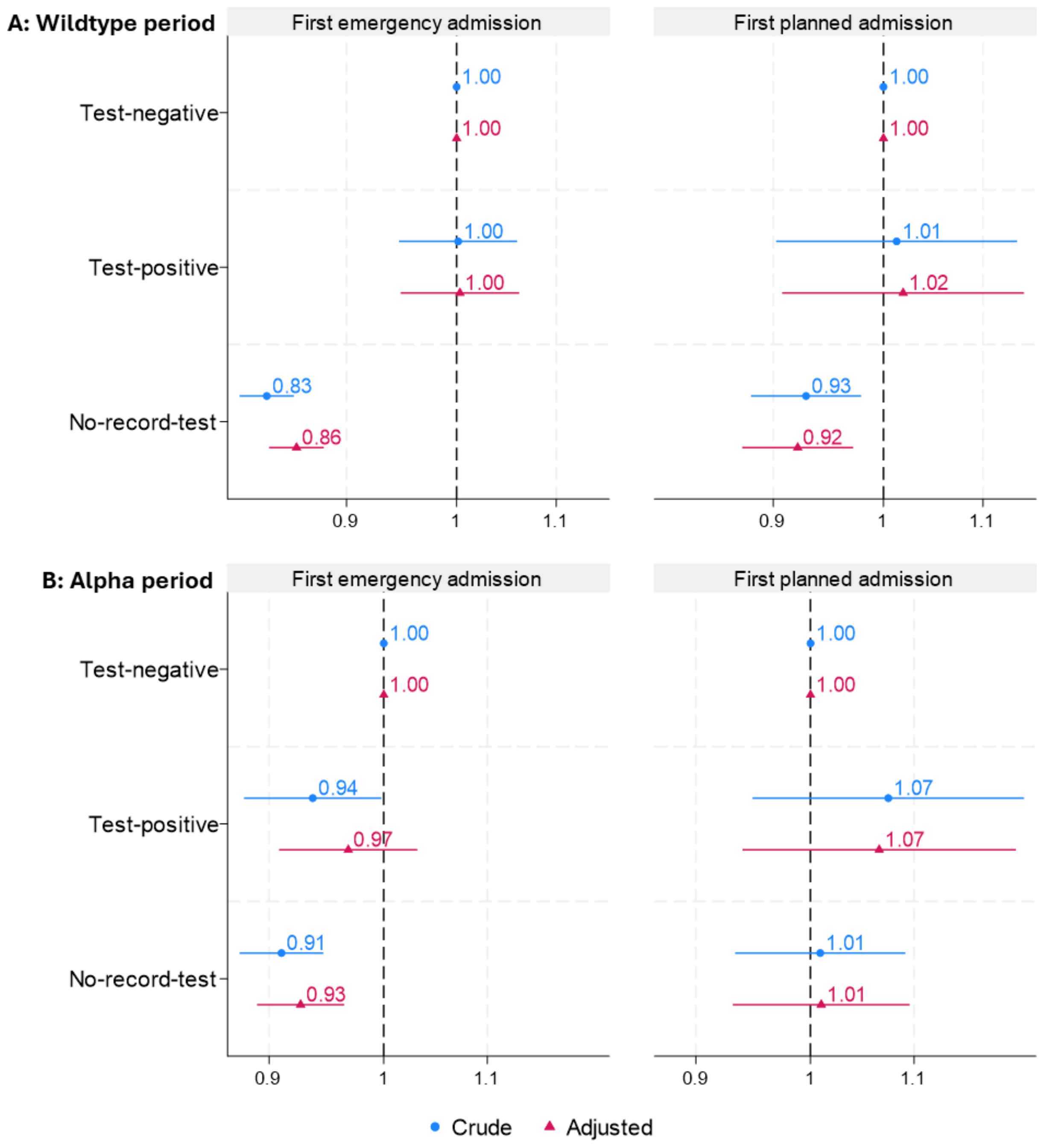


**Figure C5. Time to child's first emergency and planned admission up to 40 months old: crude and adjusted hazard ratio and 99% confidence interval by in utero SARS-CoV-2 exposure status.** In Panel A, the test-negative and test-positive groups are defined based on test results during the wildtype period, and in Panel B, based on results during the Alpha period. The no-recorded-result group is the same in both panel A and panel B and comprises children whose mother had no recorded SARS-CoV-2 test result during pregnancy in either wildtype or Alpha period. Adjusted models include maternal ethnicity, deprivation decile, maternal chronic conditions, maternal age, London residence, and month of conception.

**Table C1. Admission rates per 1,000 child-years (99% CI) by primary diagnosis, including all emergency and planned admissions up to 40 months old. The test-negative and positive groups under the "wild-type period" and "Alpha period" columns are defined based on test results during the wild-type period and Alpha period, respectively. The no-recorded-result group comprises children whose mothers had no recorded SARS-CoV-2 test result during pregnancy in both the wild-type and Alpha period.**

| Disease group | Number of disease cases | Number of admissions | Wild-type period | | Alpha period | | No-recorded-result | Total |
|---|---|---|---|---|---|---|---|---|
| | | | Test-negative | Positive | Test-negative | Positive | | |
| **Certain infectious and parasitic diseases** | | | | | | | | |
| Bacterial infectious diseases | 7,692 | 8,880 | 11.9 (11.0,12.7) | 14.1 (12.2,16.2) | 11.4 (10.2,12.7) | 9.9 (8.4,11.6) | 9.8 (9.5,10.1) | 10.2 (9.9,10.5) |
| Non-bacterial infectious diseases | 13,955 | 19,337 | 25.1 (23.9,26.4) | 27.6 (24.9,30.6) | 21.4 (19.8,23.2) | 20.7 (18.6,23.2) | 21.7 (21.2,22.1) | 22.2 (21.8,22.6) |
| **Neoplasms** | | | | | | | | |
| Malignant neoplasms | 193 | 4,420 | 5.7 (5.2,6.4) | 11.0 (9.3,12.9) | 5.1 (4.4,6.1) | -# | 5.0 (4.8,5.2) | 5.1 (4.9,5.3) |
| Benign neoplasms | 906 | 1,501 | 2.7 (2.3,3.2) | 1.1 (0.7,1.8) | 1.7 (1.3,2.3) | 1.9 (1.3,2.8) | 1.6 (1.5,1.7) | 1.7 (1.6,1.8) |
| **Diseases of the blood and blood-forming organs and certain disorders involving the immune mechanism** | 806 | 2,502 | 2.2 (1.9,2.6) | 4.4 (3.4,5.7) | 0.99 (0.7,1.4) | 2.8 (2.0,3.7) | 3.0 (2.9,3.2) | 2.9 (2.7,3.0) |
| **Endocrine, nutritional and metabolic diseases** | 1,228 | 2,373 | 3.3 (2.9,3.88) | 2.2 (1.5,3.1) | 2.4 (1.9,3.0) | 2.7 (1.9,3.6) | 2.7 (2.5,2.8) | 2.7 (2.6,2.9) |
| **Mental and behavioural disorders** | 330 | 360 | 0.4 (0.3,0.6) | 0.2 (0.0,0.6) | 0.4 (0.2,0.7) | 0.3 (0.1,0.7) | 0.4 (0.4,0.5) | 0.4 (0.4,0.5) |
| **Diseases of the nervous system** | 1,902 | 3,435 | 4.5 (4.0,5.1) | 5.0 (4.0,6.4) | 3.8 (3.1,4.6) | 3.8 (3.0,5.0) | 3.9 (3.7,4.1) | 3.9 (3.8,4.1) |
| **Diseases of the eye and adnexa** | 1,596 | 1,907 | 2.4 (2.1,2.9) | 2.6 (1.9,3.7) | 2.2 (1.7,2.8) | 2.0 (1.4,2.8) | 2.1 (2.0,2.3) | 2.2 (2.1,2.3) |
| **Diseases of the ear and mastoid process** | 2,258 | 2,626 | 3.9 (3.4,4.4) | 3.9 (3.0,5.2) | 3.2 (2.6,3.9) | 2.7 (1.9,3.6) | 2.9 (2.7,3.0) | 3.0 (2.9,3.2) |
| **Diseases of the circulatory system** | | | | | | | | |
| Heart diseases | 268 | 461 | 0.5 (0.4,0.8) | 0.4 (0.2,0.9) | 0.5 (0.3, 0.9) | 0.3 (0.1,0.8) | 0.5 (0.5,0.6) | 0.5 (0.5,0.6) |
| Circulatory diseases | 290 | 358 | 0.5 (0.4,0.7) | 0.4 (0.2,0.9) | 0.5 (0.3, 0.8) | 0.6 (0.3,1.1) | 0.4 (0.3,0.4) | 0.4 (0.4,0.5) |
| **Diseases of the respiratory system** | 31,816 | 45,898 | 66.5 (64.5,68.5) | 59.7 (55.7,64.0) | 59.3 (56.5, 62.2) | 55.3 (51.6,59.1) | 50.0 (49.3,50.7) | 52.7 (52.1, 53.3) |
| **Diseases of the digestive system** | 7,534 | 9,762 | 12.9 (12.0,13.8) | 12.8 (11.1,14.9) | 11.6 (10.4,13.0) | 10.8 (9.2,12.6) | 10.9 (10.6,11.2) | 11.2 (10.9,11.5) |
| **Diseases of the skin and subcutaneous tissue** | 3,330 | 3,954 | 4.9 (4.3,5.4) | 5.7 (4.6,7.2) | 4.3 (3.6,5.1) | 4.5 (3.5,5.7) | 4.5 (4.3,4.7) | 4.5 (4.4,4.7) |
| **Diseases of the musculoskeletal system and connective tissue** | 805 | 1,040 | 1.1 (0.8,1.4) | 1.3 (0.8,2.1) | 1.1 (0.8,1.5) | 1.5 (1.0,2.3) | 1.2 (1.1,1.3) | 1.2 (1.1,1.3) |
| **Diseases of the genitourinary system** | 3,482 | 5,436 | 6.5 (5.9,7.1) | 6.0 (4.8,7.5) | 5.6 (4.8, 6.6) | 5.6 (4.6,7.0) | 6.3 (6.0,6.5) | 6.2 (6.0,6.5) |
| **Certain conditions originating in the perinatal period** | 19,576 | 22,541 | 27.7 (26.4,29.1) | 28.2 (25.5,31.2) | 25.9 (24.1, 27.8) | 24.5 (22.1,27.1) | 25.4 (24.9,25.9) | 25.8 (25.3,26.2) |
| **Congenital malformation, deformations and chromosomal abnormalities** | 7,247 | 11,366 | 14.3 (13.4,15.3) | 12.8 (11.1,14.9) | 12.6 (11.4,14.0) | 12.1 (10.4,13.9) | 12.9 (12.6,13.3) | 13.0 (12.7,13.4) |
| **Injury, poisoning, and certain other consequences of external causes** | 8,608 | 9,990 | 11.9 (11.0,12.8) | 14.3 (12.4,16.5) | 10.8 (9.6,12.0) | 10.2 (8.7,12.0) | 11.4(11.1,11.7) | 11.5(11.2,11.8) |
| **Total** | 95,715 | 197,735 | 261.7 (257.7,265.8) | 263.9 (255.3,272.8) | 234.0 (228.4,239.8) | 215.0 (207.9,222.7) | 220.8 (219.3,222.2) | 226.8 (225.5,228.1) |

### Supplementary STROBE Checklist

STROBE Statement—Checklist of items that should be included in reports of ***cohort studies***

| | Item No | Recommendation | Page No |
|---|---|---|---|
| **Title and abstract** | 1 | (*a*) Indicate the study's design with a commonly used term in the title or the abstract<br>(*b*) Provide in the abstract an informative and balanced summary of what was done and what was found | 1-2 |
| **Introduction** | | | |
| Background/ rationale | 2 | Explain the scientific background and rationale for the investigation being reported | 3-4 |
| Objectives | 3 | State specific objectives, including any prespecified hypotheses | 4 |
| **Methods** | | | |
| Study design | 4 | Present key elements of study design early in the paper | 5 |
| Setting | 5 | Describe the setting, locations, and relevant dates, including periods of recruitment, exposure, follow-up, and data collection | 5-6 |
| Participants | 6 | (*a*) Give the eligibility criteria, and the sources and methods of selection of participants. Describe methods of follow-up<br>(*b*) For matched studies, give matching criteria and number of exposed and unexposed | 5-6 |
| Variables | 7 | Clearly define all outcomes, exposures, predictors, potential confounders, and effect modifiers. Give diagnostic criteria, if applicable | 7-8 |
| Data sources/ measurement | 8* | For each variable of interest, give sources of data and details of methods of assessment (measurement). Describe comparability of assessment methods if there is more than one group | 4-8 |
| Bias | 9 | Describe any efforts to address potential sources of bias | 5-9 |
| Study size | 10 | Explain how the study size was arrived at | 9-10 |
| Quantitative variables | 11 | Explain how quantitative variables were handled in the analyses. If applicable, describe which groupings were chosen and why | 5-8 |
| Statistical methods | 12 | (*a*) Describe all statistical methods, including those used to control for confounding<br>(*b*) Describe any methods used to examine subgroups and interactions<br>(*c*) Explain how missing data were addressed<br>(*d*) If applicable, explain how loss to follow-up was addressed<br>(*e*) Describe any sensitivity analyses | 7-9 |
| **Results** | | | |

| | | | |
|---|---|---|---|
| Participants | 13* | (a) Report numbers of individuals at each stage of study—eg numbers potentially eligible, examined for eligibility, confirmed eligible, included in the study, completing follow-up, and analysed<br>(b) Give reasons for non-participation at each stage<br>(c) Consider use of a flow diagram | 9-10 |
| Descriptive data | 14* | (a) Give characteristics of study participants (eg demographic, clinical, social) and information on exposures and potential confounders<br>(b) Indicate number of participants with missing data for each variable of interest<br>(c) Summarise follow-up time (eg, average and total amount) | 10-12 |
| Outcome data | 15* | Report numbers of outcome events or summary measures over time | 10-13 |
| Main results | 16 | (*a*) Give unadjusted estimates and, if applicable, confounder-adjusted estimates and their precision (eg, 95% confidence interval). Make clear which confounders were adjusted for and why they were included<br>(*b*) Report category boundaries when continuous variables were categorized<br>(*c*) If relevant, consider translating estimates of relative risk into absolute risk for a meaningful time period | 13-16 |
| Other analyses | 17 | Report other analyses done—eg analyses of subgroups and interactions, and sensitivity analyses | 17 |
| **Discussion** | | | |
| Key results | 18 | Summarise key results with reference to study objectives | 17 |
| Limitations | 19 | Discuss limitations of the study, taking into account sources of potential bias or imprecision. Discuss both direction and magnitude of any potential bias | 17-18 |
| Interpretation | 20 | Give a cautious overall interpretation of results considering objectives, limitations, multiplicity of analyses, results from similar studies, and other relevant evidence | 18-20 |
| Generalisability | 21 | Discuss the generalisability (external validity) of the study results | 17-18 |
| **Other information** | | | |
| Funding | 22 | Give the source of funding and the role of the funders for the present study and, if applicable, for the original study on which the present article is based | 22 |

*Give information separately for exposed and unexposed groups.

**Note:** An Explanation and Elaboration article discusses each checklist item and gives methodological background and published examples of transparent reporting. The STROBE checklist is best used in conjunction with this article (freely available on the Web sites of PLoS Medicine at http://www.plosmedicine.org/, Annals of Internal Medicine at http://www.annals.org/, and Epidemiology at http://www.epidem.com/). Information on the STROBE Initiative is available at http://www.strobe-statement.org.

18. Charlson Comorbidity Index: A Critical Review of Clinimetric Properties | Psychotherapy and Psychosomatics | Karger Publishers. https://karger.com/pps/article/91/1/8/826493/Charlson-Comorbidity-Index-A-Critical-Review-of.

19. English indices of deprivation 2019. *GOV.UK* https://www.gov.uk/government/statistics/english-indices-of-deprivation-2019.

20. Statistical geographies - Office for National Statistics. https://www.ons.gov.uk/methodology/geography/ukgeographies/statisticalgeographies.

21. Hviid, M. M., Skovlund, C. W., Mørch, L. S. & Lidegaard, Ø. Maternal age and child morbidity: A Danish national cohort study. *PLOS ONE* **12**, e0174770 (2017).

22. Hardelid, P. *et al.* SARS-CoV-2 tests, confirmed infections and COVID-19-related hospital admissions in children and young people: birth cohort study. *bmjpo* **6**, e001545 (2022).
